# Investigating the health effects of 3 coexisting tobacco and nicotine products using system dynamics population modeling: An Italian population case study

**DOI:** 10.1101/2021.04.12.21255338

**Authors:** Oscar M Camacho, Andrew Hill, Stacy Fiebelkorn, Aaron Williams, James J Murphy

**Affiliations:** British American Tobacco, R&D, Regents Park Road, Southampton, Hampshire, SO15 8TL, UK; Ventana Systems UK Ltd, Alexandra House, St Johns Street, Salisbury, SP1 2SB, UK

**Keywords:** Tobacco, Nicotine, e-cigarettes, Tobacco Heating Products, Population Modeling

## Abstract

With the proliferation of tobacco and nicotine products, there might be a need for more complex models than current two-product models. We have developed a three-product model able to represent interactions between 3 products in the marketplace. We also investigate if using several implementations of two-product models could provide sufficient information to assess 3 coexisting products. Italy is used as case-study with THPs and e-cigarettes as the products under investigation. We use transitions rates estimated for THPs in Japan and e-cigarettes in the USA to project what could happen if the Italian population were to behave as the Japanese for THP or USA for e-cigarettes. Results suggest that three-product models may be hindered by data availability while two product models could miss potential synergies between products. Both, THP and E-Cigarette scenarios, led to reduction in life-years lost although the Japanese THP scenario reductions were 3 times larger than the USA e-cigarette projections.

## Introduction

The concept of tobacco harm reduction was outlined by the Institute of Medicine (IOM) in 2001, in their monograph titled *Clearing the Smoke* (IoM, 2001). They suggested a strategy for tobacco harm reduction based on the replacement of risky tobacco products, particularly conventional cigarettes with lower risk products that provide consumers with a satisfactory alternative to conventional cigarettes. Products with potential for harm reduction were defined as products able to reduce users’ exposure to one or more tobacco toxicants with these exposure reductions having a disease relevant impact.

Since then, several tobacco and nicotine products have been developed around the world that have been suggested as such potential reduced risk products (PRRP) and have been assessed for their role in the tobacco harm reduction paradigm. U.S. Food and Drug Administration (FDA) suggested in their Modified Risk Tobacco Products (MRTP) application guidance (MRTPA, 2012) to use mathematical models, as part of a weight of evidence approach, to evaluate the potential population health effects from launching new tobacco products in the USA. A diverse range of models have been developed for this purpose, many of them conducted and funded by tobacco product manufacturers as part of their MRTP submission strategies, but this topic has also attracted academic attention (Lee et al. 2021). All these models investigate the effect of launching a new, potentially reduced risk tobacco or nicotine product with respect to a scenario where only conventional cigarettes are available (Bachand et al 2017, Djurdjevic et al 2018, Levy et al 2017). However, given the success of product categories such as e-cigarettes or tobacco heated products (THP) in markets around the world, a two-product model may not be sufficient to appropriately represent tobacco and nicotine product use dynamics. For example, in a market with already a PRRP and conventional cigarettes, introducing a new tobacco product with a risk higher than the already marketed PRRP could lead to increased population harm if the appeal of the new product causes established users of the already marketed PRRP (rather than smokers) to switch to the new PRRP,. In this paper we consider whether a three-product model would be necessary in that situation or whether evaluation of two independent two-product models could be a satisfactory approach to investigate the potential population health effects from launching a product that perturbs the dynamics affecting a PRRP already in the marketplace.

Briefly, THPs are an emerging category of tobacco products in many markets and they have proved highly successful in Asian markets like Japan and South Korea (Hori et al, 2020). THPs have been shown to produce fewer toxicants than conventional cigarettes because the tobacco is heated without combustion, leading to reduced exposure to toxicants in clinical studies and therefore, they are expected to have a lower-risk profile than cigarettes (Forster et al 2017, Gale et al 2018, Schaller et al 2016). Around the world, e-cigarettes are more widespread than THPs. In 2018 in the USA, 3.2% of all adults reported to be current users and 14.9% ever users (Villarroel et al 2020). Commonly, e-cigarettes do not contain tobacco and are comprised of a battery connected to a heating element and an e-liquid container. Although there are a wide range of products within this category, in general, the aerosol formed by e-cigarettes has been reported to be simpler and with lower level of toxicants than cigarette smoke and people using e-cigarettes exclusively are exposed to lower level of toxicants than smokers (Margham et al 2016, Chen et al 2017).

There are currently few markets where e-cigarettes and THPs coexist, but Italy was one of the first markets to experience this with THPs launched in 2014 and vaping products already available for nearly a decade (Gallus et al, 2014), but still, very little is known about the potential population health effects of these products in Italy (Lui et al. 2020). Manufactured cigarettes were the dominant product in Italy in 2012 (Gallus, et al, 2013). Two years later, 0.4% of people aged 15+ were regular igarette users. This increased to 1.8% in 2016 and then declined to 1.3% in 2018. THP products have also increased in market share with reports suggesting an increase from 0.01% in 2015 to 0.67% in 2017 and this share is expected to continue growing (Liu et al, 2018). There is no information about transitions between the various products, but Italy has excellent smoking and demographic data easily accessible from Italian public sources that make it a good candidate for modelling purposes.

Based on a previous two-product model (Hill et al. 2018), we have developed a three-product model able to assess interaction between three products. We describe its complexity and the vast amount of data required to inform such a model. Then, using our two-product model, we assess whether two independent implementations of a two-product model could be used to investigate potential health outcomes and hence avoid the complexity of a three-product model. To illustrate the different modelling approaches, we use e-cigarettes and THPs as the two coexisting PRRPs and Italy as the population of interest.

## Methods

### Product use definitions

All the product modes distinguish the analysed population through individuals’ smoking characteristics. The basis for defining a person by their smoking characteristics in this study was determined by the data source used for smoking prevalence, in this case the ISTAT reporting of the “Multipurpose survey on households: aspects of daily life”. The survey definition of a Current Smoker was someone who answered yes to the question “*did they currently smoke*?” The definition of a Former Smoker is someone that replied no to the same question, but in a follow up question responded that “*they used to smoke in the past”*. A Never Smoker is anyone that does not fall into either of those two categories and an Ever Smoker is the combined grouping of Current and Former Smokers.

### Models description

Our System Dynamics population models have been ensembled sequentially from simpler models to represent either one, two or three tobacco products in a marketplace.

All models are run from 2001 to 2100, using a time step of 1 year.

Prior to the introduction of new tobacco products to the marketplace, the one product stock and flow model of a population characterised by their cigarette smoking behaviour is shown in Figure 1

**Figure 1.**
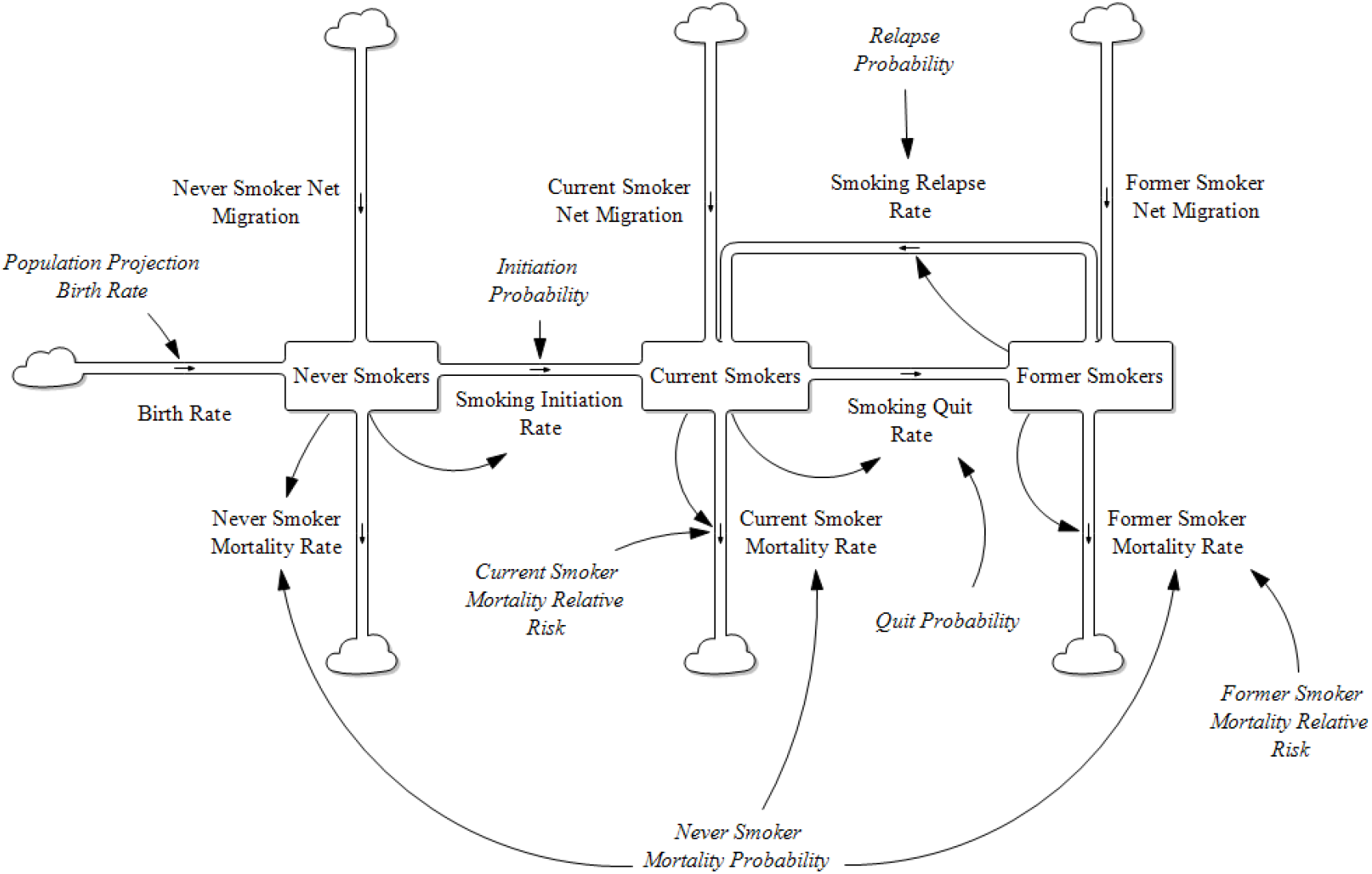
One Product Model Stock and Flow Diagram

All the stocks and flows are further categorised by gender and single year age cohorts. The Former Smoker stock is further broken down by the number of years abstinent from smoking up to 20 years and then a combined grouping of 20+ years.

The model tracks population sizes by gender and single year ages (0 – 100). At each time step, populations are aged by one year less any mortality rate at that age. The mortality rate for those aged 100 is set to be 100%.

The introduction of a single PRRP such as e-cigarettes to the marketplace requires an expanded two-product model. The model’s complexity increases reflecting the incorporation of the smoking status groups created with the number of tobacco characterised stocks increasing from 3 to 9 and potential product use transitions to 27 (Figure 2). In a one product model there is only one potential product transition flow from each population stock (initiate, quit or relapse), in the two-product model there are three potential transition outflows from each population stock.

**Figure 2.**
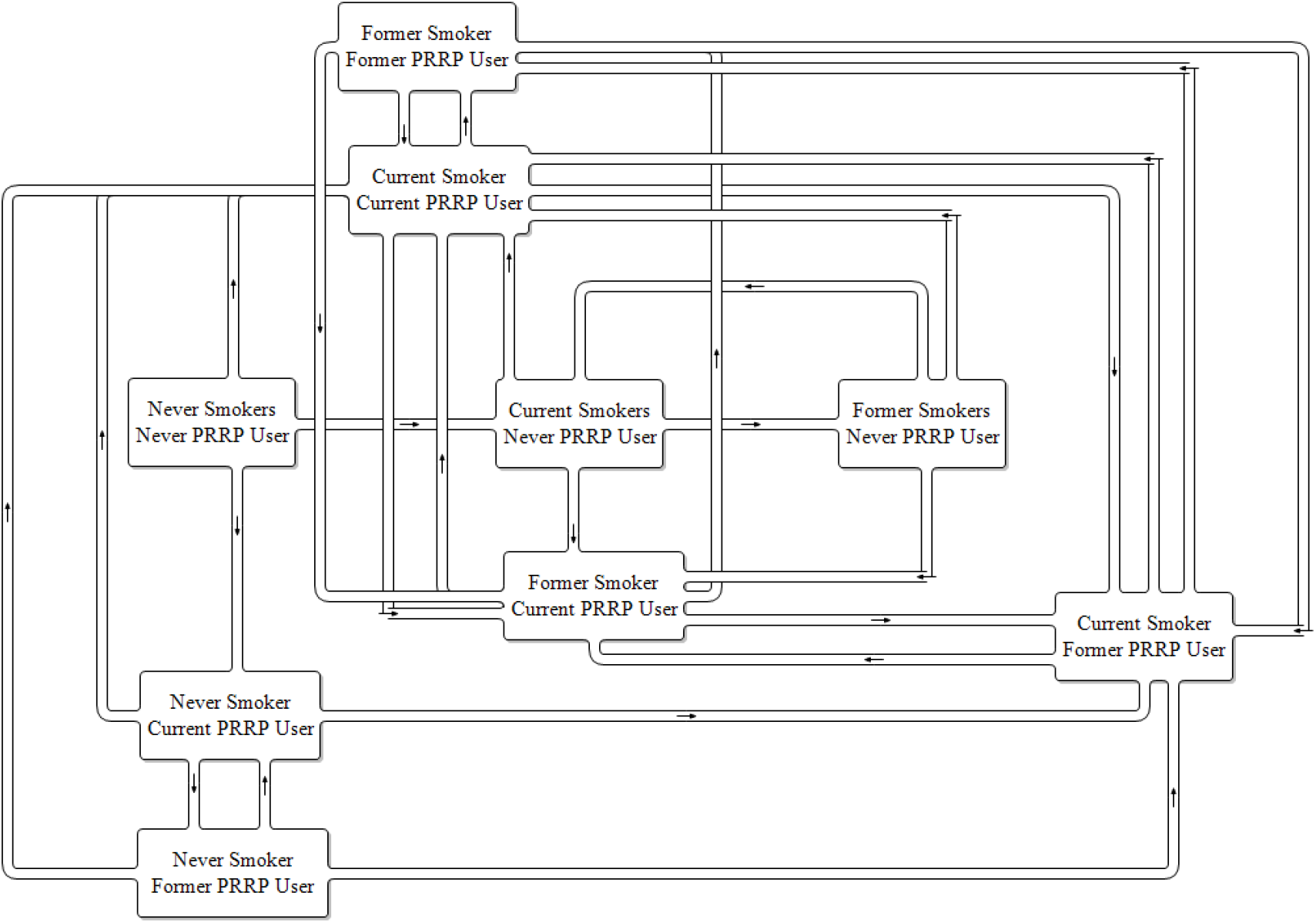
Two Product Model Stock and Flow Diagram. PRRP – Potentially Reduced Risk Product

Based on previous models it is easy to see how a three-product model expands the model structural complexity further still. In a market where there are two PRRPs available, the three-product model expands the number of stocks of population tobacco product usage further to 27 each with 7 potential product use transitions from each, a total of 189 potential transition flows.

The models were developed in Vensim DSS® software. The number of variables by type for the two and three product models are shown in Table 1.

**Table 1.** Model Type Variables

| Model | Levels | Initial | Constant | Data | Flow | Aux | Lookup | Subscripts |
| --- | --- | --- | --- | --- | --- | --- | --- | --- |
| 2 Product | 34 | 45 | 61 | 4 | 15 | 300 | 4 | 28 |
| 3 Product | 151 | 296 | 67 | 3 | 66 | 1169 | 0 | 128 |

With subscripting included, the two-product model has 48 thousand equations calculated at each time step, the three-product model has 104 thousand.

As data regarding transitions for e-cigarettes, THPs and interactions between the three-products were not available, in its place best “guess” estimates based on smoking have been used. Details about the data inputs and multiple assumptions required to inform the three-product model can be found in ***Supplementary File 1***. For the two-product model implementations, in order to provide more useful projections than modellers’ best estimates, we used published transition data between smoking and e-cigarettes estimated for the USA population (Brouwer et al, 2020) and transitions between THP and smoking estimated for the Japanese population (Camacho et al, 2021). In the E-Cigarette scenarios, e-cigarettes are considered to be in the Italian market from 2010 while THP products are introduced in scenarios from 2014. More details about the data inputs for the two-product models can be found in Table 2 while, a description of the two-product model formulation and data initialisation is contained in ***Supplementary File 2***. Model calibration and verification is provided in ***Supplementary File 3***.

**Table 2.**
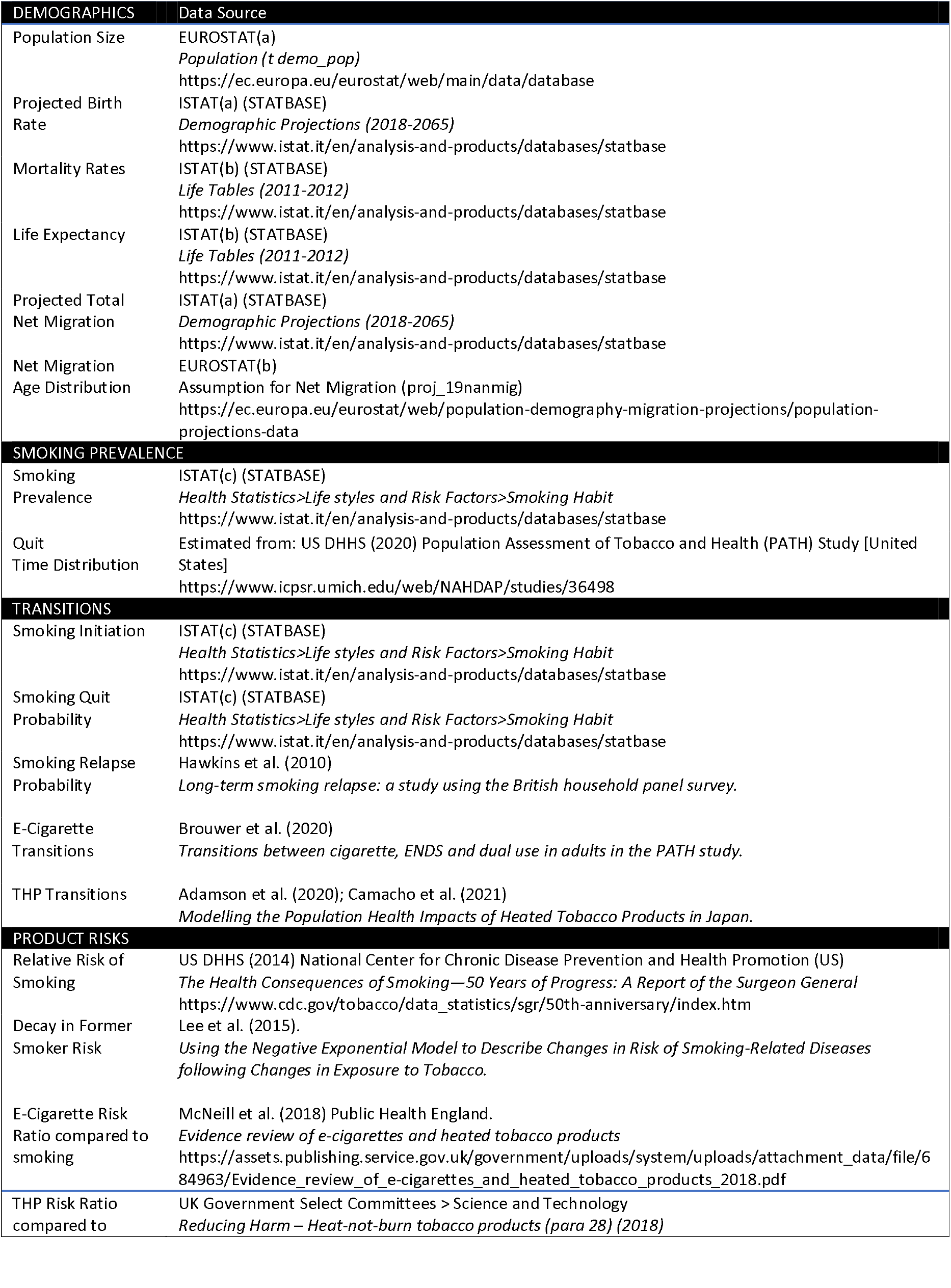
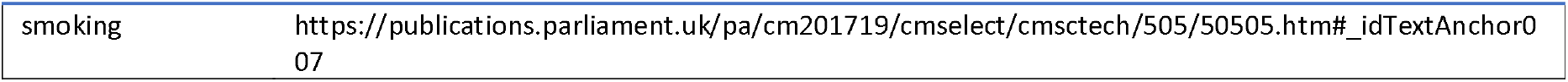
Two-Product Model Data Sources

## Analyses

Scenarios for our three-product model are built to illustrate data requirements and outputs.

### Three-product model scenarios

Two main scenarios were produced for illustrative purpose only displaying comparisons in terms of life-years lost cumulatively up to the year 2100. The scenarios were:

1. Baseline scenario in which neither e-cigarettes or THPs have ever existed.
2. Scenario representing current Italian market with only e-cigarettes and conventional cigarettes.
3. Scenario representing current Italian market with both e-cigarettes and THP products. This scenario itself was assessed under 6 different conditions. Scenario 3.1 considered that initiation of e-cigarettes and THP use are mutually exclusive or initiation between those groups are not competitive. Then we also assessed what would be the life-years projections if different percentages (20% −100%) of e-cigarette users (lowest risk product) were to instead initiate THP use.

### Two-product model scenarios

The projection scenarios assessed are described below. Information within brackets denotes whether the scenario was modelled using mean estimates of transition probability (*deterministic*) or from random sampling the estimate from with 95% confidence interval ranges (*sensitivity*).

#### Scenario Descriptions

1. **Smoking Only (Deterministic)** The model includes cigarettes as the only tobacco product available.
2. **Smoking and E-Cigarettes (Deterministic)** E-cigarettes are added as the only alternative product to cigarettes in 2010.
3. **Smoking and THP (Deterministic)** THPs are added as the only alternative product to cigarettes in 2014.
4. **Combined E-Cigarette and THPs at a ratio of 50:50 (Deterministic)** A single pseudo PRRP is generated by combining e-cigarette & THP transition rates at 50% of each. In addition, the risk ratio of the pseudo PRRP is set to the midpoint between the e-cigarette and THP risk ratio, 7.5% that of combustible cigarettes.
5. **Combined E-Cigarette and THPs at a ratio of 63:37 (Deterministic)** E-cigarette & THP transition rates are combined at 63% of e-cigarettes and 37% THPs values. In Japan, where both PRRP products are established, a nationwide population survey estimated the patterns of use of tobacco product use in adolescents (Kuwabara et al. 2020). A simple ratio of ever use of either PRRP product is used to generate the 63:37 ratio. In addition, the risk ratio of the pseudo PRRP is set to the weighted ratio of the e-cigarette and THP risk, 6.85% that of combustible cigarettes.
6. **Smoking and E-Cigarettes Transitions (Sensitivity)** E-cigarettes are added as the only alternative product to cigarettes in 2010 with transition probabilities selected from within the 95% confidence interval range using random selection routine to generate one thousand sensitivity runs.
7. **Smoking and THP (Transitions (Sensitivity)** THPs are added as the only alternative product to cigarettes in 2014 with transition probabilities selected from within the 95% confidence interval range using random selection routine to generate one thousand sensitivity runs.

Additionally, we repeated scenarios 1-5 with a new “what if” assumption:

a. Smoking initiation rates decline at 3% per year from 2020 onwards.

## Results

### Three-product model

For illustrative purposes only, when comparing life-years lost in Scenario 1 (cigarettes only) and Scenario 2 (two products available, cigarettes and e-cigarettes), the model indicates that life years lost due to smoking could be reduced by 17.6 million by 2100 (Table 3).

**Table 3.** Reduction in Life-years lost by 2100 when comparing Scenario 2 (cigarettes and e-cigarettes available) and scenario 3 (all three products) to Scenario 1 (only cigarettes available).

| Scenarios<br>(Reduction in million Life-years lost) | 2010 | 2020 | 2030 | 2040 | 2050 | 2060 | 2070 | 2080 | 2090 | 2100 |
| --- | --- | --- | --- | --- | --- | --- | --- | --- | --- | --- |
| 2. Only e-cigarettes | 0.0 | 0.07 | 0.57 | 1.70 | 3.36 | 5.33 | 7.51 | 10.15 | 13.79 | 17.61 |
| 3. Both PRRPs | 0.0 | 0.07 | 0.72 | 2.59 | 5.65 | 9.46 | 13.71 | 18.63 | 24.82 | 31.03 |
| 3.1 20% vapers initiate THP | 0.0 | 0.07 | 0.72 | 2.59 | 5.65 | 9.47 | 13.70 | 18.62 | 24.79 | 30.99 |
| 3.2 40% vapers initiate THP | 0.0 | 0.07 | 0.72 | 2.59 | 5.65 | 9.47 | 13.69 | 18.60 | 24.74 | 30.94 |
| 3.3 80% vapers initiate THP | 0.0 | 0.07 | 0.72 | 2.59 | 5.65 | 9.47 | 13.67 | 18.50 | 24.60 | 30.77 |
| 3.4 All vapers initiate THP | 0.0 | 0.07 | 0.72 | 2.59 | 5.65 | 9.47 | 13.63 | 18.38 | 24.42 | 30.56 |

Compared with Scenario 3, Scenario 3 (all three products are available) is estimated to lead to an additional reduction of 13.4m life-years lost by 2100. Thus, with THPs and e-cigarettes available, the total reduction of life-years lost would be 31 million. In other words, introducing both products would have a synergistic effect that increases the potential overall benefit. Table 3 also shows projections of what would happen if e-cigarette users were to initiate into THP use instead of vaping at different rates, i.e., initiating e-cigarettes with a risk ratio of 10% rather than one with 5%.

### Two-product models

In Table, the smoking initiation and cessation probabilities are static over the projection period. The projected smoking prevalence in people aged 14 and over reduces from 19.7% (2010) to 11.3% (2100) a reduction of 8.4%. The fall in prevalence occurs mostly in the period up to 2050 (8.1%).

Based on Japan transition data, the THP scenario projects the greatest drop in smoking prevalence, with 4.7% smoking prevalence by the end of the projection period. In comparison, the inclusion of e-cigarettes, based on USA transitions, only reduces smoking prevalence down to 9.6%. The scenarios with a mix of e-cigarettes and THPs project slightly lower smoking prevalence at 8.2% (50/50 mix) and 8.7% (63/37 mix).

E-cigarette prevalence, when introduced alongside smoking, peaks at 2.7% in 2020 and then declines to a plateau of 1.6% before starting to rise again slightly in 2100. THP prevalence, when introduced alongside smoking, peaks at 9.6% in 2030 and then declines to a plateau of 7.1%. The scenarios using a mix of both e-cigarettes and THPs are above those with e-cigarettes only and below those with THPs only.

Comparing the reduction of life years lost across alternative scenarios against those projected for the smoking only scenario, the largest health gain is 10.7m for the THP only scenario. This is over three times the number for the E-Cigarette only scenario, which projects a reduction of 3.0m live years lost. Taking a similar approach as per the three-product model, if all these e-cigarette users were using THP (with risk ratio of 10%) the new projection for life years lost would be 2.9m.

To this point all projections were based on fixed smoking initiation probabilities. In Table 5 the same scenarios assessed before are drawn with 3% annual reduction in the smoking initiation. These alternative scenarios provide a more realistic representation in many markets with smoking declines independently of any PRRP introduced to the market. Under these conditions, the Smoking Only scenario projected that smoking prevalence in people aged 14 and over reduces from 19.7% (2010) to 2.9% (2100), a reduction of 16.8 percentage points. The fall in smoking prevalence occurs more rapidly in earlier projection years but, a year on year drop, is projected across all years.

**Table 4.** Projected prevalence and reduction of life years lost for Scenarios 1-5

| Scenarios 1- 5 | 2010 | 2020 | 2030 | 2040 | 2050 | 2060 | 2070 | 2080 | 2090 | 2100 |
| --- | --- | --- | --- | --- | --- | --- | --- | --- | --- | --- |
| **Smoking Prevalence (%)** |  |  |  |  |  |  |  |  |  |  |
| Smoking Only | 19.7 | 16.6 | 14.3 | 12.7 | 11.6 | 11.2 | 11.3 | 11.2 | 11.2 | 11.3 |
| Smoking + E-Cigarettes | 19.7 | 15.3 | 12.7 | 10.9 | 9.8 | 9.5 | 9.5 | 9.5 | 9.5 | 9.6 |
| Smoking + THPs | 19.7 | 13.7 | 7.3 | 5.4 | 4.7 | 4.6 | 4.6 | 4.6 | 4.6 | 4.7 |
| Smoking + 50% E-Cig 50% THPs | 19.7 | 14.5 | 11.1 | 9.3 | 8.4 | 8.1 | 8.1 | 8.1 | 8.1 | 8.2 |
| Smoking + 63% E-Cig 37% THPs | 19.7 | 14.8 | 11.7 | 9.9 | 8.9 | 8.6 | 8.6 | 8.6 | 8.6 | 8.7 |
| **PRRP Prevalence (%)** |  |  |  |  |  |  |  |  |  |  |
| Smoking Only | 0.0 | 0.0 | 0.0 | 0.0 | 0.0 | 0.0 | 0.0 | 0.0 | 0.0 | 0.0 |
| Smoking + E-Cigarettes | 0.0 | 2.7 | 2.2 | 1.8 | 1.6 | 1.6 | 1.6 | 1.6 | 1.6 | 1.7 |
| Smoking + THPs | 0.0 | 6.7 | 9.6 | 8.9 | 7.8 | 7.3 | 7.2 | 7.1 | 7.1 | 7.1 |
| Smoking + 50% E-Cig 50% THPs | 0.0 | 4.6 | 4.0 | 3.1 | 2.6 | 2.5 | 2.5 | 2.5 | 2.5 | 2.6 |
| Smoking + 63% E-Cig 37% THPs | 0.0 | 4.0 | 3.3 | 2.6 | 2.2 | 2.2 | 2.2 | 2.2 | 2.2 | 2.2 |
| **Reduction of Life Years Lost Compared to Smoking Only Scenario (Million Person Years)** |  |  |  |  |  |  |  |  |  |  |
| Smoking Only | 0.0 | 0.0 | 0.0 | 0.0 | 0.0 | 0.0 | 0.0 | 0.0 | 0.0 | 0.0 |
| Smoking + E-Cigarettes | 0.0 | 0.0 | 0.2 | 0.6 | 1.2 | 1.7 | 2.1 | 2.5 | 2.7 | 3.0 |
| Smoking + THPs | 0.0 | 0.0 | 0.7 | 2.3 | 4.4 | 6.2 | 7.6 | 8.8 | 9.8 | 10.7 |
| Smoking + 50% E-Cig 50% THPs | 0.0 | 0.0 | 0.4 | 1.1 | 2.1 | 2.9 | 3.7 | 4.3 | 4.8 | 5.2 |
| Smoking + 63% E-Cig 37% THPs | 0.0 | 0.0 | 0.3 | 0.9 | 1.7 | 2.4 | 3.0 | 3.5 | 4.0 | 4.3 |

**Table 5.** Projected prevalence and reduction of life years lost for Scenarios 1a-5a

| Scenarios 1a- 5a | 2010 | 2020 | 2030 | 2040 | 2050 | 2060 | 2070 | 2080 | 2090 | 2100 |
| --- | --- | --- | --- | --- | --- | --- | --- | --- | --- | --- |
| **Smoking Prevalence (%)** |  |  |  |  |  |  |  |  |  |  |
| Smoking Only | 19.7 | 16.6 | 14.0 | 11.6 | 9.4 | 7.7 | 6.3 | 5.0 | 3.8 | 2.9 |
| Smoking + E-Cigarettes Only | 19.7 | 15.3 | 12.4 | 9.9 | 7.9 | 6.4 | 5.1 | 3.9 | 2.7 | 1.7 |
| Smoking + THPs Only | 19.7 | 13.7 | 7.1 | 4.5 | 3.3 | 2.5 | 2.0 | 1.5 | 1.1 | 0.8 |
| Smoking + 50% E-Cig 50% THPs | 19.7 | 14.5 | 10.8 | 8.4 | 6.6 | 5.3 | 4.3 | 3.4 | 2.5 | 1.8 |
| Smoking + 63% E-Cig 37% THPs | 19.7 | 14.8 | 11.4 | 8.9 | 7.1 | 5.7 | 4.6 | 3.6 | 2.7 | 1.9 |
| **PRRP Prevalence (%)** |  |  |  |  |  |  |  |  |  |  |
| Smoking Only | 0.0 | 0.0 | 0.0 | 0.0 | 0.0 | 0.0 | 0.0 | 0.0 | 0.0 | 0.0 |
| Smoking + E-Cigarettes Only | 0.0 | 2.7 | 2.2 | 1.7 | 1.5 | 1.3 | 1.2 | 1.1 | 1.0 | 1.0 |
| Smoking + THPs Only | 0.0 | 6.7 | 9.6 | 8.5 | 6.9 | 5.6 | 4.7 | 3.9 | 3.3 | 2.8 |
| Smoking + 50% E-Cig 50% THPs | 0.0 | 4.6 | 4.0 | 2.9 | 2.2 | 1.9 | 1.7 | 1.5 | 1.3 | 1.2 |
| Smoking + 63% E-Cig 37% THPs | 0.0 | 4.0 | 3.3 | 2.4 | 1.9 | 1.7 | 1.5 | 1.3 | 1.2 | 1.1 |
| **Reductions in Life Years Lost Compared to Smoking Only Scenario (Million Person Years)** |  |  |  |  |  |  |  |  |  |  |
| Smoking Only | 0.0 | 0.0 | 0.0 | 0.0 | 0.0 | 0.0 | 0.0 | 0.0 | 0.0 | 0.0 |
| Smoking + E-Cigarettes Only | 0.0 | 0.0 | 0.2 | 0.6 | 1.2 | 1.7 | 2.1 | 2.4 | 2.5 | 2.6 |
| Smoking + THPs Only | 0.0 | 0.0 | 0.7 | 2.3 | 4.4 | 6.1 | 7.5 | 8.5 | 9.3 | 9.9 |
| Smoking + 50% E-Cig 50% THPs | 0.0 | 0.0 | 0.4 | 1.1 | 2.1 | 2.9 | 3.6 | 4.1 | 4.5 | 4.7 |
| Smoking + 63% E-Cig 37% THPs | 0.0 | 0.0 | 0.3 | 0.9 | 1.7 | 2.4 | 3.0 | 3.4 | 3.7 | 3.9 |

After introduction of THPs, the scenario projects the greatest drop in smoking prevalence, with 0.8% smoking prevalence by the end of the projection period. In comparison, the inclusion of e-cigarettes reduces smoking prevalence down to 1.7%. The scenarios with a mix of e-cigarettes and THPs project smoking prevalence’s of 1.8% (50/50 mix) and 1.9% (63/37 mix).

E-cigarette prevalence, when introduced alongside smoking, peaks at 2.7% in 2020 and then declines to a plateau of 1.0%. This value is lower than from the scenario with static initiation probabilities as expected, as there will be fewer smokers to transition to e-cigarettes. THP prevalence, when introduced alongside smoking, peaks at 9.6% in 2030 and then declines to 2.8% by 2100. The projections from scenarios using a mix of both e-cigarettes and THPs are above those for e-cigarettes only and below those for THPs only.

Comparing to the smoking only scenario, the largest reduction in life years lost is 9.9m for the THP only scenario, down from 10.7m previously projected when the 3% annual decline was not applied to the initiation rate. However, comparatively, reductions in projections are still over three times the life years lost projected using the E-Cigarette only scenario, which projects a reduction of 2.6m in life years lost.

### Sensitivity Assessment

Sensitivity assessments present 1000 projections drawn using the transitions’ 95% confidence intervals from either the E-Cigarette scenario based on USA data (Brouwer et al, 2020) and THP scenario based on Japan data (Camacho et al, 2021). Reductions in life years lost (as life years saved for brevity) for the E-Cigarette and THP scenarios are displayed in figures 3 and 4. Additional graphs reporting other aspects of the sensitivity assessment can be found in ***Supplementary File 3***.

**Figure 3.**
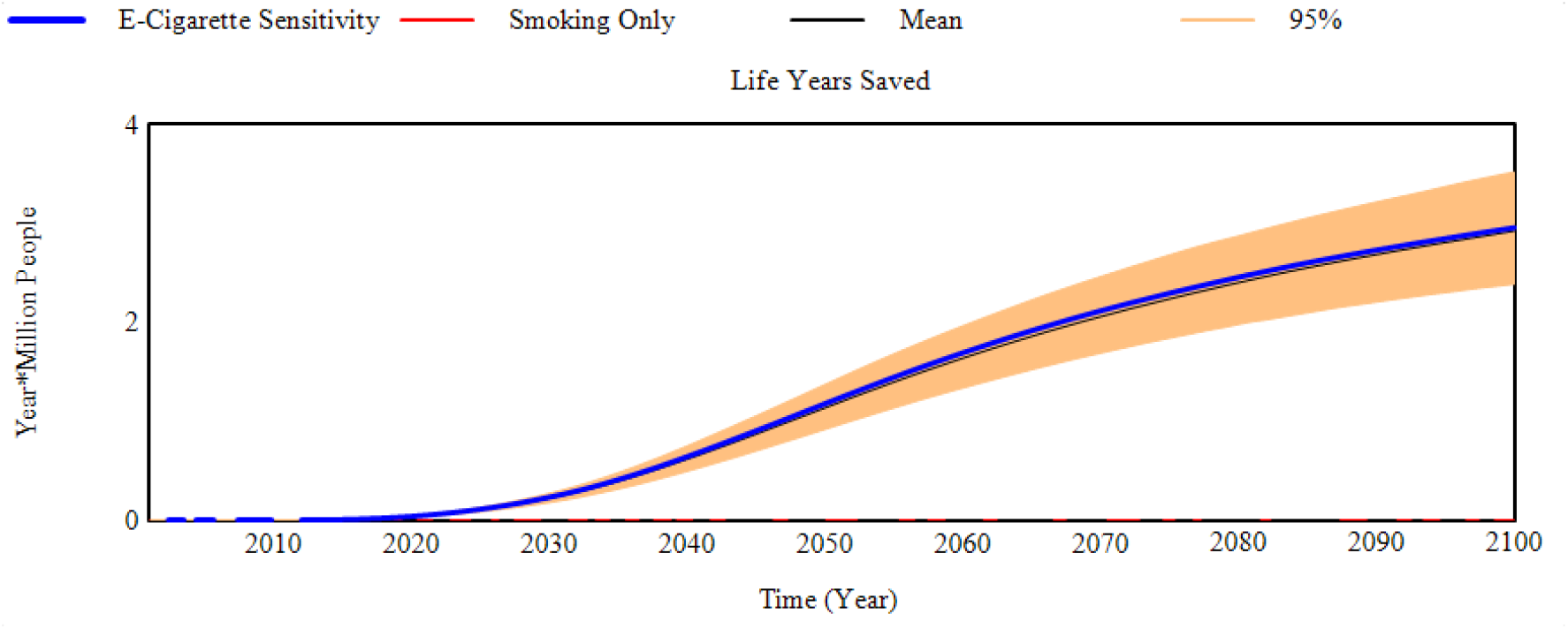
Reduction in life years lost compared to the smoking only scenario (Life years saved) for the E-Cigarette scenario sensitivity to transition probabilities.

**Figure 3.**
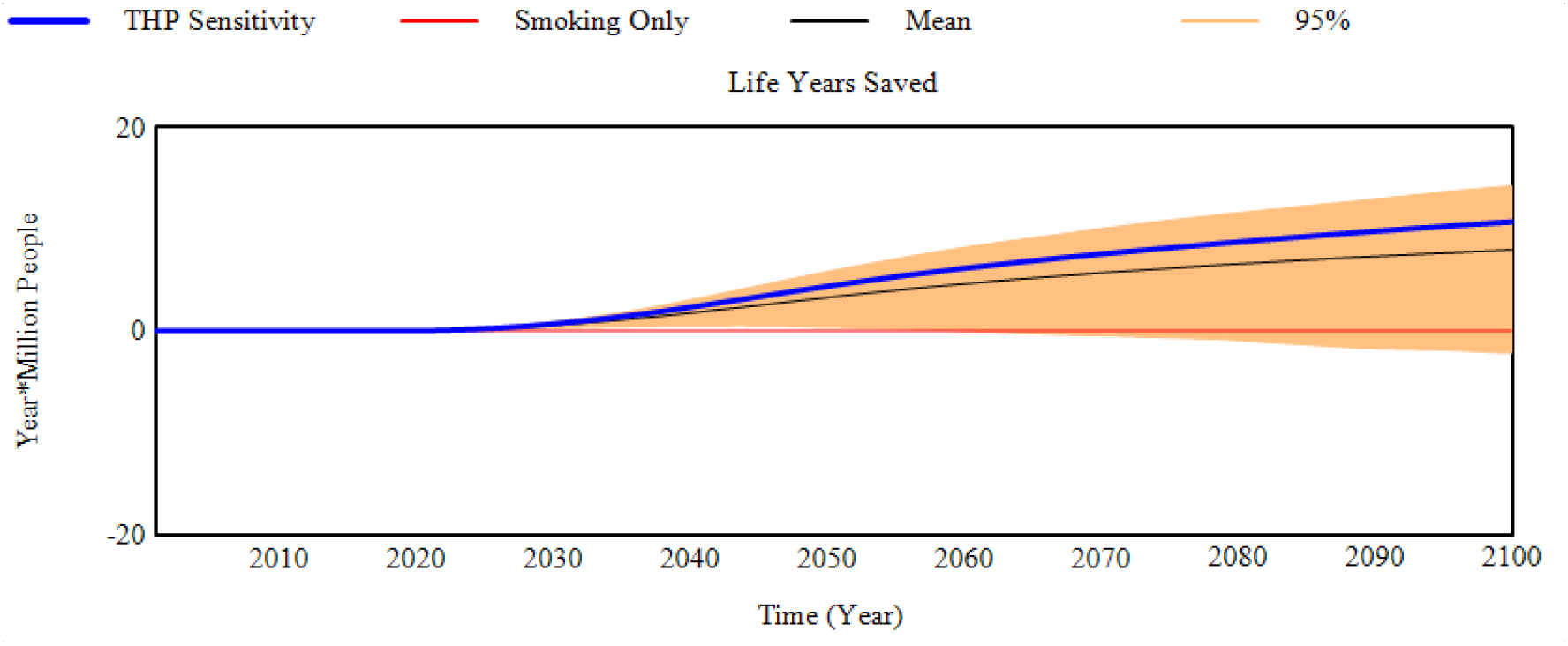
Reduction in life years lost compared to the smoking only scenario (Life years saved) for the THP scenario sensitivity to transition probabilities.

The model projections were not overly sensitivity to randomly selecting combinations of e-cigarette transition probabilities from within their 95% confidence inter range (Table 6). By comparison, the model projections were more sensitive to the random selection of THP transition probabilities from within their confidence intervals. This is most prominent in the projection of reduction in life years lost. Although the mean and median values (8.0m and 8.7m respectively) were positive in terms of reducing life years lost compared to a smoking only scenario, certain randomly selected THP transition probabilities generated greater life years lost when compared to the smoking only scenario. Approximately 5% of the 1000 sensitivity runs suggest an increase in life years lost (Table 7), with the maximum of life years lost across all 1000 scenarios at 5.8m of additional life years that could be lost.

**Table 6.** Summary Sensitivity Results across all 1000 projections for the E-Cigarette and THP scenarios

| Sensitivity Results | Min | Max | Mean | Median | Standard Deviation |
| --- | --- | --- | --- | --- | --- |
| **E-Cigarette Transition Sensitivity** |  |  |  |  |  |
| Smoking Prevalence (%) | 9.1 | 10.1 | 9.6 | 9.6 | 0.2 |
| PRRP Prevalence (%) | 1.4 | 1.8 | 1.5 | 1.5 | 0.1 |
| Life Years Saved (M) | 2.0 | 3.9 | 2.9 | 2.9 | 0.3 |
| **THP Transition Sensitivity** |  |  |  |  |  |
| Smoking Prevalence (%) | 2.9 | 11.2 | 5.5 | 5.4 | 1.3 |
| PRRP Prevalence (%) | 2.5 | 21.0 | 6.5 | 5.7 | 2.8 |
| Reduction in Life Years Lost (M) | -5.8 | 15.7 | 8.0 | 8.7 | 4.2 |

**Table 7.**
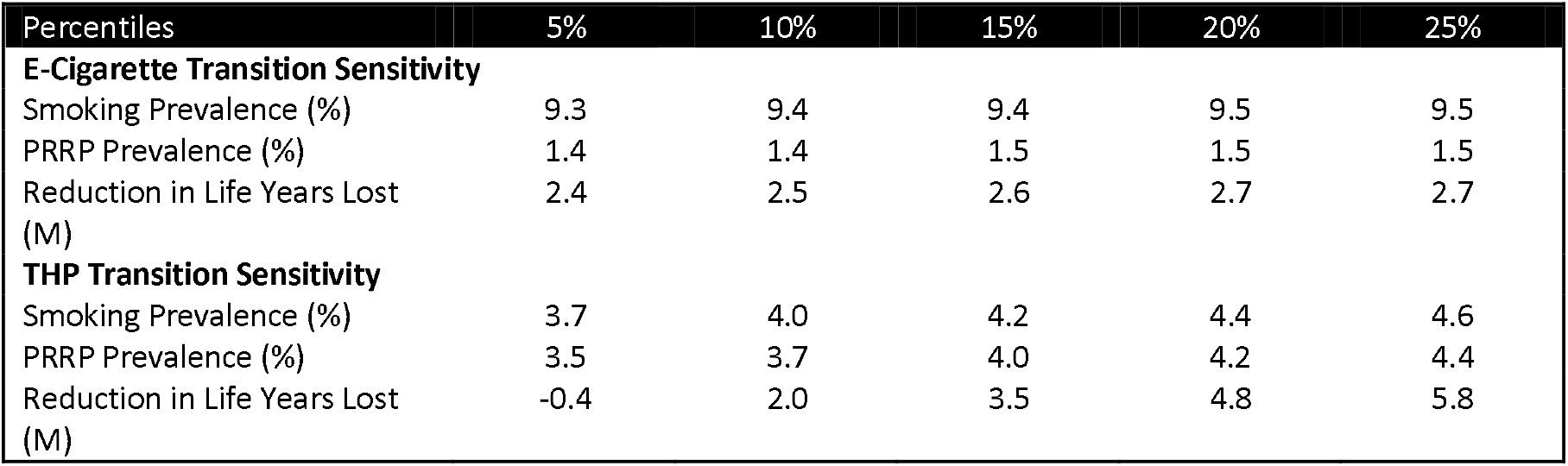
Sensitivity Percentiles for all 1000 projections for the E-Cigarette and THP scenarios

| Percentiles | 5% | 10% | 15% | 20% | 25% |
| --- | --- | --- | --- | --- | --- |
| **E-Cigarette Transition Sensitivity** |  |  |  |  |  |
| Smoking Prevalence (%) | 9.3 | 9.4 | 9.4 | 9.5 | 9.5 |
| PRRP Prevalence (%) | 1.4 | 1.4 | 1.5 | 1.5 | 1.5 |
| Reduction in Life Years Lost (M) | 2.4 | 2.5 | 2.6 | 2.7 | 2.7 |
| **THP Transition Sensitivity** |  |  |  |  |  |
| Smoking Prevalence (%) | 3.7 | 4.0 | 4.2 | 4.4 | 4.6 |
| PRRP Prevalence (%) | 3.5 | 3.7 | 4.0 | 4.2 | 4.4 |
| Reduction in Life Years Lost (M) | -0.4 | 2.0 | 3.5 | 4.8 | 5.8 |

It is worth noticing that some of the THP transitions in Japan study have wide 95% confidence intervals (Supplementary File 2 - **Error! Reference source not found**.), particularly from the transitions from THP Users to Dual Use. The mean probability is 3.0%, but the 95% confidence interval on this estimate ranges from 0.0% to 20.5%. These wide confidence intervals have significant leverage on the sensitivity results.

## Discussion

The use of mathematical models has been suggested to assess the potential population health effects of launching new nicotine products (MRTPA, 2012). Numerous models have been developed with this purpose (Lee et al. 2021). However, to our knowledge, all these models are two-product models and with increasing market complexity with competing PRRPs like e-cigarettes and THPs, two-product models may not be able to represent population dynamics appropriately.

We have developed a three-product model with a structure considering interactions between all three products and allowing for 27 tobacco and nicotine product use statuses. Our example illustrates how a three-product model is able to draw projections accounting for potential synergies between products that enhance harm reduction character of PRRPs. This aspect is lost in two-product models, however if the aim of the modelling assessment is to investigate the potential for a new product to introduce harm compared to a scenario where another PRRP already exists, separate models for the current and prospective PRRPs could be informative in relation to whether the prospective PRRP is likely to introduce benefit or burden at the population level.

We used the Italian population to assess e-cigarettes and THPs potential for harm reduction separately and considering that e-cigarettes were introduced in Italy earlier than THPs. Given that Italian estimates on transitions to and from these PRRPs are not available, estimates calculated based on USA behaviours were used for the e-Cigarette scenario and for the THP scenario these were Japanese estimates.

To aid interpretability, it is essential to clarify that none these models forecast future tobacco and nicotine product prevalence nor associated mortality rates. They generate projections based on data currently available and the underlying assumption that currently observed behaviour will continue. Evaluation of whether models suggest mainly positive or negative effects are informative, especially when performing sensitivity testing, as well as the magnitude of changes between different scenarios. Sensitivity testing of model input data helps to identify key data, assisting the design of future surveys to gather that information.

Limitations on data availability, especially on transitions between two differing types of PRRPs, precluded any detailed projections using the three-product model. Using the two-product model, our projections from both models suggested beneficial effects from launching either product, the THP-scenario suggested the largest point estimate in reductions of life-years lost while with higher uncertainty in outcomes. A 5% of projections in the sensitivity testing of the THP scenario pointed to additional life years lost, this was driven by wide confidence intervals for some transition estimates.

The fact that the THP scenario suggest three times greater effects reducing life years lost than the e-cigarette model only can be surprising to many given that THPs have a higher risk ratio than e-cigarettes. This effect is due to the different dynamics observed between the Japan and USA populations. One of the judgements to be assessed on all PRRPs is the balance between moving existing smokers to a less harmful product against introducing a product with some risk to people that may never have smoked and eventually leading them to smoking, known as the gateway effect.

In our scenarios, the THP scenario performed better than the e-cigarette scenario because THP initiation by never smokers in Japan is a quarter of the initiation probability of e-cigarettes in the USA and THPs in Japan have double the transition probability of shifting existing smokers to the PRRP than e-cigarettes in the USA. A beneficial effect persists despite overall greater tobacco and nicotine product use prevalence in the THP scenario and even with a higher relative risk ratio than e-cigarettes.

Independent scenarios do not provide much information about a potential combined effect from availability of both PRRPs. In order to mitigate this problem, we suggest creating combined or hybrid two-product scenarios. These combined scenarios could be designed using weights based on expected prevalence ratio for the two PRRPs placing each transition point estimate between the two estimates for the independent PRRP transitions, as well as using their confidence intervals for sensitivity testing. In our examples we used 50:50 and 63:37 ratios and when rates are considered constant over time, projections from these scenarios lay between the E-Cigarette and THP scenarios. However, this may not always be case with the non-linear relationships within system dynamic models. This effect can be observed for the scenarios with 3% annual reduction in initiation rates, where results from these scenarios suggest that the combination of different tobacco transitions could lead to a smoking prevalence outcome outside the range of solely e-cigarettes or solely THPs. For most of the projection period the combined scenario projections fall between the two sole product scenarios prevalence, but post 2090 both combined scenarios exceeded the smoking prevalence of the E-Cigarette or THP scenarios.

In Japan, nicotine containing e-cigarettes are banned and the new generation of THPs was approved only relatively recently following a marketing authorisation in the USA. Perhaps the same behaviours would have been observed in Japan if e-cigarettes were available instead THPs. Therefore, we do not know whether the alternative product would have similar transitions or not if both products coexisted in the same market under the same cultural and regulatory conditions. Thus, our results can only be interpreted as possible projections for the Italian population if they were to behave as the Japanese are using THP products or USA population with e-cigarettes. Transitions cannot be regarded purely as a product effect, as cultural, social and regulatory aspects will play an important role on transitions.

In principle, the three-product model is thought to be best suited to model the tobacco and nicotine product use behaviour of three coexisting products, especially to estimate potential synergistic effects that could be observed if the two PRRP products were to attract a different pool of smokers. However, a clear limitation of a three-product model is the type and amount of data required to appropriately inform the model, with the many transitions involving the two PRRPs not likely to be available. This will be even more problematic as a larger number of PRRPs are included in the assessment, as the number of transitions and nicotine use statuses will increase in a multiplicative manner. Alternatively, we have suggested using several two-product models, building combined scenarios to draw credible scenarios with the limitation of missing any synergistic effect.

Other general limitations of our modelling come from using transition data from different sources which leads to inconsistencies in product use definitions. Smoking data in ISTAT does not have specific time constraints for current or former smokers as they are only asked if they currently smoked or they smoked in the past. The definition of product use status for e-cigarettes are based on usage in the previous 30 days while the e-cigarette transition probabilities are estimated from a Markov multistate transition model based on data approximately at one-year intervals (Brouwer et al, 2020). The THP transition probabilities (Camacho et al, 2021) are also based on one-year intervals with product status definitions according to use in the last year. For e-cigarette initiation, this could mean that short-term experimental use of an e-cigarette may be overlooked. The authors judge that any health impact from a short-term, less than a year, e-cigarette use would be minimal.

In conclusion, we have developed a three-product model able to evaluate the population health effects of three coexisting tobacco and nicotine products and showed that different implementations of two-product models can be informative when evaluating three products. In our case study, both THPs and e-cigarettes in Italy would suggest significant reductions in life years lost if the Italian population were to behave like the Japanese population with THPs and USA population with e-cigarettes, with significantly more gains from the behaviours observed in the Japanese population.

## Supporting information

3-Product Model Formulation and Initialisation.pdf

2-Product Model Formulation and Initialisation.pdf

2-product Model Verification Final.pdf

## Data Availability

All data is available in the supplementary files.

## Abbreviations

BAT: British American Tobacco
BMJ: British Medical Journal
COT: Committee of Toxicology
ENDS: Electronic Nicotine Devices
FDA: U.S. Food and Drug Administration
ISTAT: Italian National Institute of Statistics
MAE: Mean Absolute Error
MAEoM: Mean Absolute Error Over Mean
PHE: Public Health England
PRRP: Potential Reduced Risk Product THP Tobacco Heated Product
US DHHS: U.S. Department of Health and Human Services

## Acknowledgments

A special thanks to all reviewers who have helped to add clarity to this manuscript. We would like to thank Kantar Health for their work collecting and processing the Japanese data.

## Author Contributions

OMC.: Conceptualization-Equal, Investigation-Equal, Methodology-Equal, Project administration-Lead, Supervision-Lead, Writing-original draft-Lead. AH.: Conceptualization-Equal, Data curation-Equal, Formal analysis-Lead, Methodology-Equal, Software-Lead, Writing-review & editing-Equal. SF.: Conceptualization-Supporting, Data curation-Equal, Investigation-Supporting, Project administration-Supporting, Writing-review & editing-Equal. AW.: Resources-Supporting, Writing-review & editing-Equal. JM.: Conceptualization-Equal, Funding acquisition-Lead.

