## Supplementary material for "Investigating the health effects of 3 coexisting tobacco and nicotine products using system dynamics population modeling: An Italian population case study": 3-Product Model Formulation and Initialisation.pdf

***Supplementary File 1*** **-Three-Product Model Description & Required Assumptions**

Definitions

All the product modes distinguish the population by their smoking characteristics. The basis for defining a person by their smoking characteristics is determined by the data source used for smoking prevalence, in this case the ISTAT reporting of the "Multipurpose survey on households: aspects of daily life". The survey definition of a Current Smoker was someone who answered Yes to the question "did they currently smoke?". The definition of a Former Smoker is someone that replied no to the same question, but in a follow up question responded that "they used to smoke in the past". A Never smoker is anyone that does not fall into either of those two categories and an Ever Smoker is the combined grouping of Current and Former smokers.

Three-letter reference codes for each stock were used to indicate use status (N=never used, C=current user, and F=former user) for all products in the order conventional cigarettes, e-cigarettes and THPs. Stock or product status definitions can be found in Table 1.

*Table 1. Stocks or product status and their definitions.*

| *Nicotine use status* | *Definitions* |
|---|---|
| NNN | Never smoker who has never used e-cigarettes or (Tobacco Heated Product) THPs |
| CNN | Current smoker who has never used e-cigarettes or THPs |
| NCN | Current e-cigarette user who has never smoked or used THPs |
| NNC | Current THP user who has never smoked or used e-cigarettes |
| CCN | Dual user of cigarettes and e-cigarettes who has never used THPs |
| CNC | Dual user of cigarettes and THPs who has never used e-cigarettes |
| NCC | Dual user of e-cigarettes and THPs who has never smoked |
| CCC | Current user of all three products |
| FNN | Former smoker who has never used any other product |
| NFN | Former e-cigarette user who has never used any other product |
| NNF | Former THP user who has never used any other product |
| FFN | Former smoker and e-cigarette user who has never used THPs |
| FNF | Former smoker and THPs user who has never used e-cigarettes |
| NFF | Former e-cigarettes and THP user who has never smoked |
| FFF | Former user of all three products |
| FCN | Former smoker who is currently using e-cigarettes but has never used THPs |
| FNC | Former smoker who is currently using THPs but has never used e-cigarettes |
| FCC | Former smoker who is currently using both e-cigarettes and THPs |
| CFN | Current smoker who has previously used e-cigarettes but has never used THPs |
| NFC | Current THP user who has previously used e-cigarettes but has never smoked |
| CFC | Dual user of cigarettes and THPs who has previously used e-cigarettes |
| CNF | Current smoker who has previously used THPs but has never used e-cigarettes |
| NCF | Current e-cigarette user who has previously used THPs but has never smoked |
| CCF | Dual user of cigarettes and e-cigarettes who has previously used THPs |
| FFC | Current THPs user who has previously used both cigarettes and e-cigarettes |
| FCF | Current e-cigarette user who has previously used both cigarettes and THPs |
| CFF | Current smoker who has previously used both e-cigarettes and THPs |

Three-product model data inputs and assumptions

*Assumptions to reduce model structure complexity*

As we expect some transitions would be very low in number (e.g. transitions involving a change of behaviour for all three products within the same year), to reduce unnecessary complexity and/or until more data become available, those were deemed to be 0 with exception of NNN to any other state. However, all possible states can ultimately be reached by changing two product statuses at the same time over a two-year period (e.g. CNN->FCN->CFC). Initiation of all products in a single year is possible to represent experimentation with nicotine-based products.

The maximum potential number of transitions at each time step in the model is 189 (Figure 1).

| *From* | | *To* | | | | | | |
|---|---|---|---|---|---|---|---|---|
| NNN | | CNN | NCN | NNC | CCN | CNC | NCC | CCC |
| CNN | | CCN | CNC | CCC | FNN | FCN | FNC | FCC |
| NCN | | CCN | NCC | CCC | NFN | CFN | NFC | CFC |
| NNC | | CNC | NCC | CCC | NNF | CNF | NCF | CCF |
| CCN | | CCC | FFN | FCN | FCC | CFN | CFC | FFC |
| CNC | | CCC | FNF | FNC | FCC | CNF | CCF | FCF |
| NCC | | CCC | NFF | NFC | CFC | NCF | CCF | CFF |
| CCC | | FFF | FCC | CFC | CCF | FFC | FCF | CFF |
| FNN | | CNN | CCN | CNC | CCC | FCN | FNC | FCC |
| NFN | | NCN | CCN | NCC | CCC | CFN | NFC | CFC |
| NNF | | NNC | CNC | NCC | CCC | CNF | NCF | CCF |
| FFN | | CCN | CCC | FCN | FCC | CFN | CFC | FFC |
| FNF | | CNC | CCC | FNC | FCC | CNF | CCF | FCF |
| NFF | | NCC | CCC | NFC | CFC | NCF | CCF | CFF |
| FFF | | CCC | FCC | CFC | CCF | FFC | FCF | CFF |
| FCN | | CCN | CCC | FFN | FCC | CFN | CFC | FFC |
| FNC | | CNC | CCC | FNF | FCC | CNF | CCF | FCF |
| FCC | | CCC | FFF | CFC | CCF | FFC | FCF | CFF |
| CFN | | CCN | CCC | FFN | FCN | FCC | CFC | FFC |
| NFC | | NCC | CCC | NFF | CFC | NCF | CCF | CFF |
| CFC | | CCC | FFF | FCC | CCF | FFC | FCF | CFF |
| CNF | | CNC | CCC | FNF | FNC | FCC | CCF | FCF |
| NCF | | NCC | CCC | NFF | NFC | CFC | CCF | CFF |
| CCF | | CCC | FFF | FCC | CFC | FFC | FCF | CFF |
| FFC | | CCC | FFF | FCC | CFC | CCF | FCF | CFF |
| FCF | | CCC | FFF | FCC | CFC | CCF | FFC | CFF |
| CFF | | CCC | FFF | FCC | CFC | CCF | FFC | FCF |

*Figure 1. Matrix of all possible transitions for a three-product model.*

The order of the letters represents smoking, e-cigarette use and THP use. N=never, C=current and F=former smoker/user. Each stock is subdivided by age, gender and time since quitting based on public data to enhance representation of the Italian population. The model can produce information for single-year age groups, but public data are generally reported by age categories. For the three-product model age categories were defined as <12, 12-13, 14-17, 18-19, 20-24, 25-34, 35-44, 45-54, 55-59, 60-64, 65-74 and >75 years.

For the three-product model, transition rate estimates are required not only between smoking and e-cigarettes and THPs but also between e-cigarettes and THPs. In the nicotine products risk spectrum, e-cigarette use is widely considered to be lower risk than THPs. Public health officials and regulatory authorities could be concerned about the introduction of THPs undermining e-cigarette harm reduction potential by driving e-cigarette users to THP instead.

For e-cigarette transition rates, data were scarce and had only limited availability from 2015 onwards. For THP no data could be sourced for Italy.

This required either substitution of transition rates from other geographies or assumptions made linking transition behaviour to that known for cigarette smoking. The types of assumptions that would be required in the three-product model are illustrated in Table 2. (These assumption values are from the authors only to illustrate the type of inputs required).

Table 2. Types of assumptions required for the three-Product Model.

| E-cigarette Assumptions | THP Assumptions |
|---|---|
| E-cigarette initiation is 25% of the smoking initiation probability | THP initiation is 25% of the smoking initiation probability |
| E-cigarette users have the same quit probability as smokers | THP users have the same quit probability as smokers |
| Former e-cigarette users have the same relapse probability as smokers | Former THP users have the same relapse probability as smokers |
| 80% of e-cigarette initiation would have smoked | 80% of THP initiation would have smoked |
| 1.5% of smokers switch annually to e-cigarettes | 2.5% of smokers switch annually to THP only |
| 1.5% of smokers witch to dual use with e-cigarettes | 1.5% of smokers switch annually to dual use with THP |
| 1.5% of e-cigarette users switch back to smoking | 1.5% of e-cigarette user switch annually to THP only |
| 1.5% of dual smoking/e-cigarette users switch back to smoking only | 1.5% of e-cigarette user switch annually to dual use with THP |
| 1.5% of dual smoking/e-cigarette users switch to e-cigarettes only | 1.5% of THP users switch to smoking |
| | 0.5% of THP users switch to e-cigarettes |
| | 1.5% of dual smoking/THP switch to smoking only |

Illustrative Analyses

Performing model projections under these assumptions, three main scenarios were produced to illustrate comparisons in terms of life-years lost cumulatively up to the year 2100, plus 4 additional variants of Scenario 3.

The scenarios investigated were:

1. Baseline scenario in which neither e-cigarettes or THPs have ever existed.
2. Addition of e-cigarettes to the marketplace

4. Both e-cigarettes and THP added to the marketplace (THP at 70% risk reduction relatively to smoking)
    a. 20% vapers initiate THP
    b. 40% vapers initiate THP
    c. 80% vapers initiate THP
    d. All vapers initiate THP

We also performed a sensitivity analysis to test the importance of some of the assumptions used as input values. We can investigate the tipping point for the excess risk (ER) parameter for THP under Scenario 3 assumptions (ie, at what point the ER of THP would lead to population burden in terms of mortality).

Illustrative Results

In Scenario 1, in which no new nicotine products would have been launched and only conventional cigarettes would be available in Italy, the model projects that based on the smoking trend from 2001, more than 137 million life-years would be lost by 2100 (Table 3).

Table 3. Life-years lost (millions) by 2100 under Scenario 1.

| | 2010 | 2020 | 2030 | 2040 | 2050 | 2060 | 2070 | 2080 | 2090 | 2100 |
|---|---|---|---|---|---|---|---|---|---|---|
| Scenario 1 | 16.0 | 33.8 | 51.4 | 68.1 | 83.1 | 96.0 | 107.3 | 117.8 | 127.7 | 137.1 |

With the introduction of e-cigarettes (Scenario 2), smoking prevalence is projected to decrease from 24.4% in 2001 to 12.3% by 2100 (Figure 2). Meanwhile, e-cigarette prevalence could reach 8.9% of the population by that date.

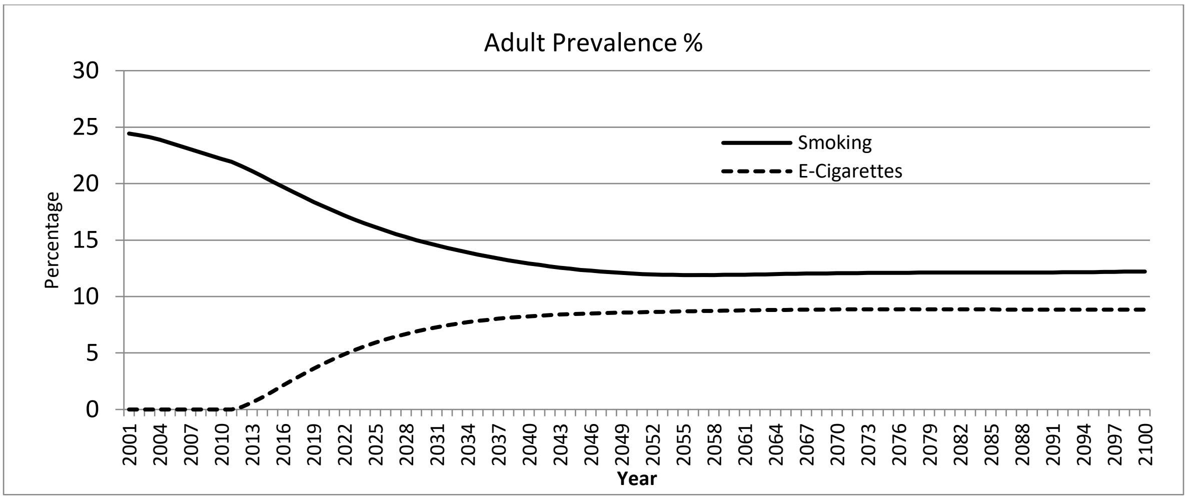


Figure 2. Prevalence projections based on scenario 2.

With Scenario 3, which includes THPs as well as e-cigarettes, smoking prevalence is estimated to reduce to 9.3% by 2100. Prevalence for e-cigarettes would be 8.9% and 10.1% for THP by the same year (Figure 3.)

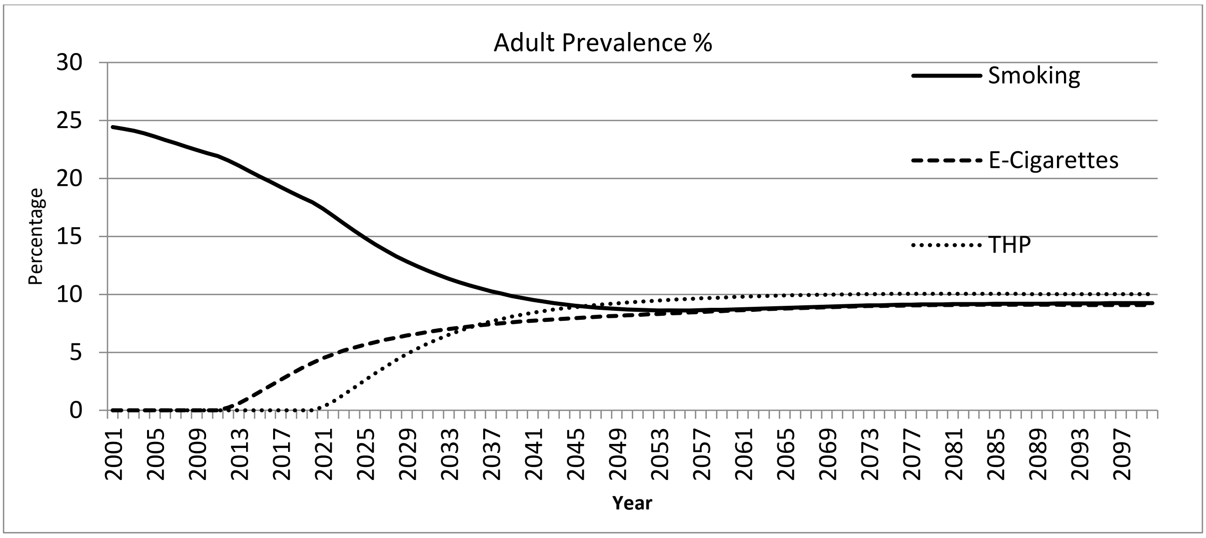


*Figure 3. Projection of smoking and product use prevalence based on scenario 3 assumptions.*

Positive trends are also suggested if a 70% in risk reduction for THPs is used (Table 4.) As expected, the 20% increase in risk in scenario 4 with respect of scenario 2 leads to a reduction of life-years saved. The model indicates that could be 20.3 million life-years saved by 2100 if 100% of vapers were to transition to THP use. This is still a more favourable outcome compared to the scenario where only e-cigarettes are available.

*Table 4. Life-years saved by 2100 comparing scenario 1 (only cigarettes available) to scenario 2 (cigarettes and e-cigarettes available) and to scenario 4 (all 3 products are available) and with THP risk reduction at 70% compared to continue smoking.*

| Scenarios<br>Reduction in Life Years Lost(M) | 2010 | 2020 | 2030 | 2040 | 2050 | 2060 | 2070 | 2080 | 2090 | 2100 |
|---|---|---|---|---|---|---|---|---|---|---|
| 2. Only e-cigarettes | 0.0 | 0.07 | 0.57 | 1.70 | 3.36 | 5.33 | 7.51 | 10.15 | 13.79 | 17.61 |
| 4. Both Products | 0.0 | 0.07 | 0.72 | 2.57 | 5.49 | 8.94 | 12.66 | 16.88 | 22.25 | 27.76 |
| 4.1 50% vapers initiate THP | 0.0 | 0.07 | 0.72 | 2.57 | 5.43 | 8.71 | 12.51 | 15.99 | 20.91 | 26.06 |
| 4.2 All vapers initiate THP | 0.0 | 0.07 | 0.72 | 2.57 | 5.29 | 8.06 | 10.54 | 13.02 | 16.38 | 20.36 |

Another type of output for the three-product model could be risk ratio tipping point assessment. For example, based on our assumptions the model suggests that THP products could have the potential for harm reduction so long as they were to reduce risk by at least 27% relative to conventional cigarettes. RRs up to 73% showed positive numbers of life-years saved by 2100 (Figure 4).

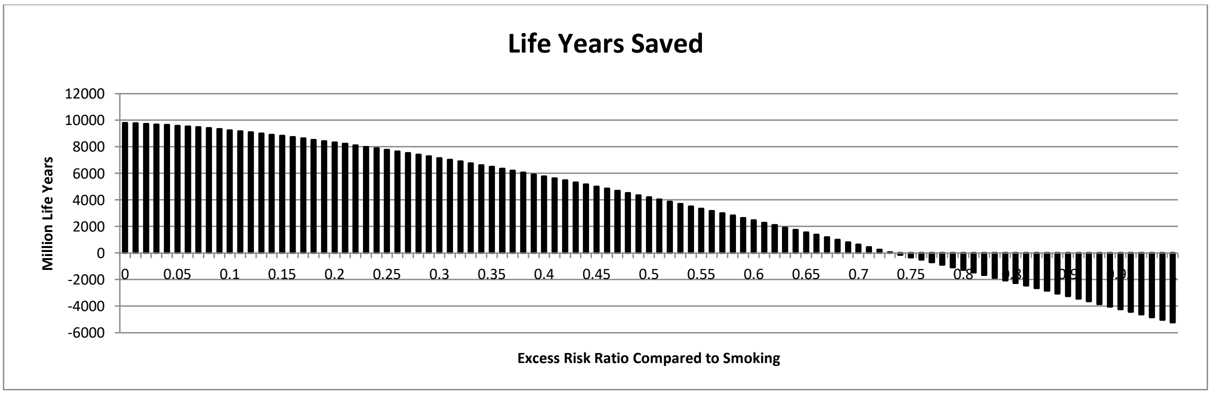


*Figure 4. Tipping point assessment of the risk ratio of THP relative to smoking.*
