## Supplementary material for "Investigating the health effects of 3 coexisting tobacco and nicotine products using system dynamics population modeling: An Italian population case study": 2-Product Model Formulation and Initialisation.pdf

*Supplementary File 2* – **Two-Product Model Description**

**Model Initialisation**

Prior to the introduction of the PRRPs, the two-product model was initialised and calibrated with cigarettes as the only tobacco product.

Each stock is initiated by taking the Italian population by each gender by age in the base year 2001 (EUROSTAT(a)) and applying the ISTAT(a) estimated smoking status prevalence at that date (Table 1). ISTAT did not provide prevalence estimates for those under 14, we assumed 100% these ages as 100% Never Smokers.

*Table 1 Initial Smoking Status Prevalence*

| | Never Smoker | | Current Smoker | | Former Smoker | |
|---|---|---|---|---|---|---|
| | Male | Female | Male | Female | Male | Female |
| Under 12s | 100.0% | 100.0% | 0.0% | 0.0% | 0.0% | 0.0% |
| 12-14 | 100.0% | 100.0% | 0.0% | 0.0% | 0.0% | 0.0% |
| 14-17 | 87.1% | 91.4% | 9.5% | 5.2% | 3.4% | 3.4% |
| 18-19 | 70.4% | 76.9% | 24.1% | 17.5% | 5.5% | 5.6% |
| 20-24 | 55.2% | 71.3% | 36.2% | 20.6% | 8.6% | 8.1% |
| 25-34 | 47.8% | 67.0% | 38.6% | 20.8% | 13.6% | 12.2% |
| 35-44 | 39.2% | 56.9% | 38.3% | 25.8% | 22.5% | 17.3% |
| 45-54 | 30.8% | 59.6% | 37.1% | 22.5% | 32.1% | 17.9% |
| 55-59 | 29.8% | 65.2% | 32.2% | 19.3% | 38.0% | 15.5% |
| 60-64 | 33.3% | 73.3% | 25.9% | 12.9% | 40.8% | 13.8% |
| 65-74 | 30.7% | 80.3% | 20.1% | 7.4% | 49.2% | 12.3% |
| 75+ | 32.7% | 86.0% | 12.4% | 3.5% | 54.9% | 10.5% |

The distribution of Former Smokers by years since quit was not available from ISTAT. We used data from The Population Assessment of Tobacco and Health (PATH) Study, a national longitudinal study of tobacco use and how it affects the health of people in the United States, to allocate former smokers to cessation length by year up to 20 years and then a combined group of 20+ years (US DHHS 2020).

The model has been developed as a closed system, with births and net migration, external model input from ISTAT(b), each drawing from a potential pool of people and mortality rates are accumulated. The birth rate is an external data input to the model and net migration is estimated as a rate per thousand current inhabitants. No data could be found to identify the smoking characteristics of the net migration flows, with net migration being allocated in proportion to the current inhabitants smoking status prevalence by age and gender.

The annual smoking initiation probabilities (Table ) and any trend in the probability are estimated separate by each gender and age by analysing the Ever Smoker prevalence between two consecutive age groups over the years 2001-2012, prior to any significant prevalence of e-cigarettes or THPs in the marketplace (ISTAT(a)). Data reported from ISTAT commenced at age 14 so any smoking initiation prior to that age is assumed into the 14 – 17 age group. Smoking initiation beyond age 34 is exceedingly small and was omitted. This is supported by the mean age for smoking initiation for men is 16.2 years (±3.1 95% CI) and 17.4 years (±3.2 95% CI) for women (Verlato et al. 2006).

*Table 2 Annual Smoking Initiation Probabilities*

| Initiation (%/year) | Under 14 | 14 – 17 Years | 18-19 Years | 20 – 24 Years | 25 – 34 Years |
|---|---|---|---|---|---|
| Male | 0.0% | 3.6% | 10.0% | 2.0% | 0.9% |
| Female | 0.0% | 3.5% | 2.7% | 1.4% | 0.3% |

Declines in smoking initiation trends in southern European countries (Italy, Portugal, Spain) stalled in the 1990s (Marcon et al. 2018). The ISTAT reporting of Ever Smoking prevalence (Figure 1) in age cohorts where smoking initiation occurred also displayed no significant trends in ages 14 to 34. As ever smoker prevalence reflects smoking initiation and is independent of quit rates, we used static smoking initiation probabilities the same as those in Table over the entire projection period 2001-2100.

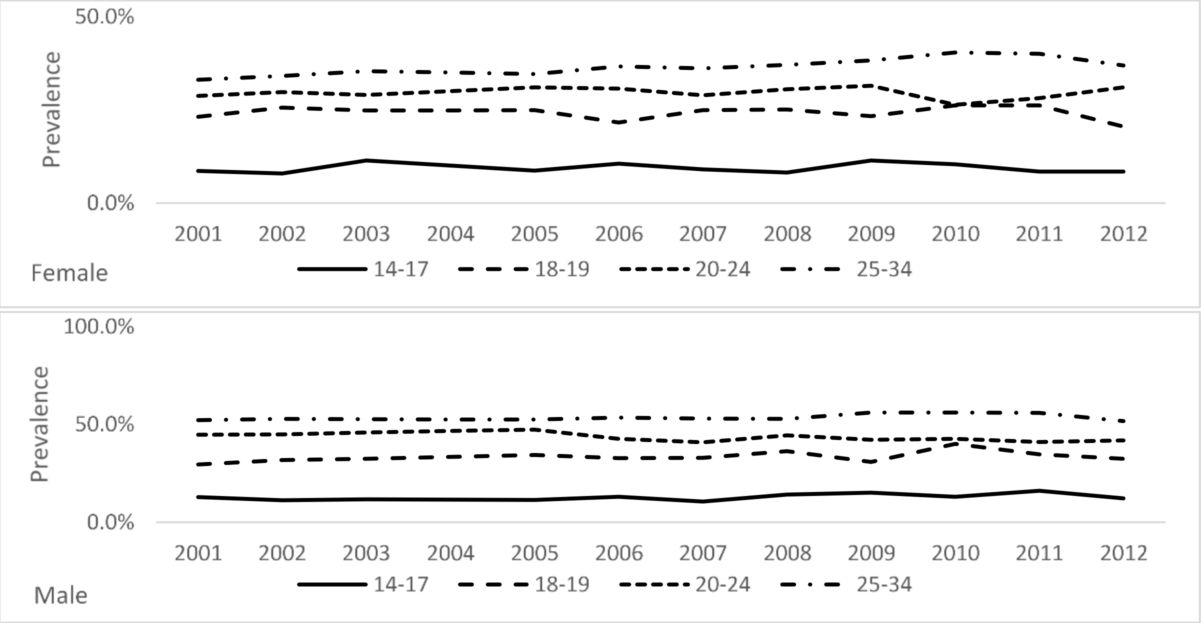


*Figure 1 Ever Smoking Prevalence*

Smoking quit probabilities are estimated by calibrating the model to observed smoking status prevalence's by gender and age cohort for the estimated initiation probabilities.

*Table 3 Annual Smoking Quit Probability*

| | 14-17 | 18-19 | 20-24 | 25-34 | 35-44 | 45-54 | 55-64 | 65-74 | 75+ |
|---|---|---|---|---|---|---|---|---|---|
| Male | 17.1% | 3.5% | 5.3% | 3.1% | 2.8% | 2.8% | 5.1% | 2.9% | 7.4% |
| Female | 25.0% | 2.8% | 10.2% | 5.0% | 1.8% | 1.3% | 2.9% | 5.2% | 3.9% |

The higher quit probabilities below the age of 18 likely reflect youth experimentation with smoking. The raised quit probability for females 20-24 (10.2%) and to a lesser extent (5.0%) at 25-34 may be a consequence of childbearing ages.

The calibration was performed over 2001-2012 (prior to any significant impact from PRRPs in the market) by adjusting the input smoking quit probabilities at the gender by age cohort level, using a method of minimising the Mean Absolute Error over Mean (MAEoM) statistic.

$$\text{MAEoM} = \frac{1}{n}\sum_{i=1}^{n}(x, - \breve{x}) \;/\; \hat{x}$$

Three separate MAEoM payoff functions were calculated, for matching Never, Current and Former prevalence's respectively, but each given equal weighting for minimising residual errors between data and model estimates.

Relapse rates are derived by the number of years a former smoker has been abstinent from smoking. Data specific to Italy could not be located. We applied data from a United Kingdom study on long term relapse rates (Hawkins et al, 2010) (Figure ). This data was not specific to gender or age.

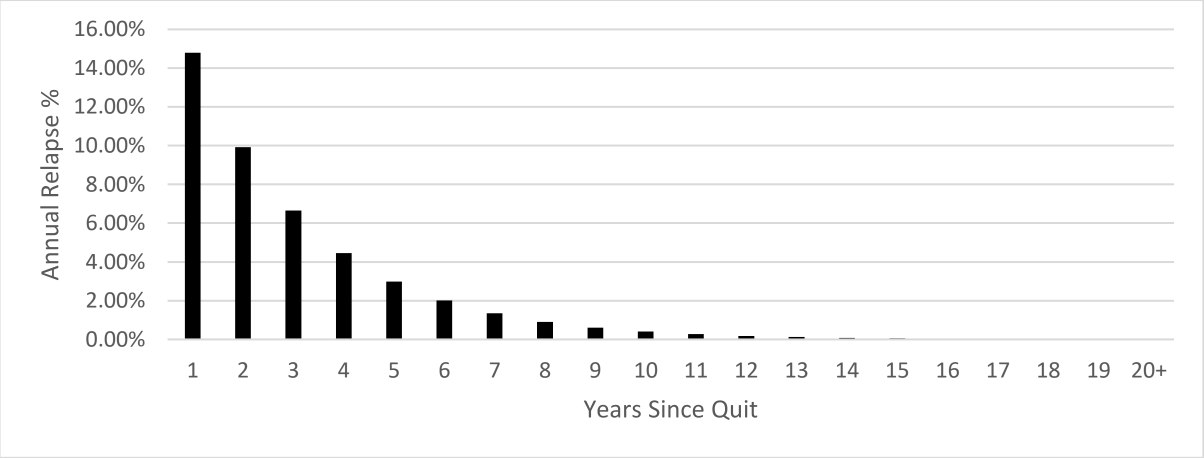


*Figure 2 Annual smoking relapse probability after years quit*

As e-cigarettes and THPs are relatively new to the marketplace there is no long-term effect data or analysis available as yet as to how quickly their risks to health will decline post cessation. We have assumed the same rate of decay of excess risk for e-cigarettes and THPs as that published for cigarette smoking (Fry et al. 2013).

Relative risk factors for all-cause mortality of each smoking status compared to a Never Smoker are taken from the Cancer Prevention Study II [US DHHS, 2014] (Table 4). These relative risks are then applied to the estimated mortality rate of a Never Smoker at each gender – age cohort level to generate mortality rates from each smoking status.

*Table 4 Relative Risk Factors*

| | 25-34 | 35-44 | 45-54 | 55-59 | 60-64 | 65-74 | 75+ |
|---|---|---|---|---|---|---|---|
| **Male** | 1 | 2.55 | 2.55 | 2.97 | 2.97 | 3.02 | 2.4 |
| **Female** | 1 | 1.79 | 1.79 | 2.63 | 2.63 | 2.87 | 2.47 |

For former smokers, the excess risk decay over time was derived using a negative exponential distribution method (Lee et al. 2015).

$$\textit{Former Smoker Excess Risk} =$$

$$\textit{Current Smoker Excess Risk(Gender,Age)} \; x \; EXP(\frac{Years\ Quit\ x\ ln\ (2)}{Half\ Life\ Decay\ Period})$$

$$\textit{Former Smoker Relative Risk} = \textit{Former Smoker Excess Risk} + 1$$

For scenarios with PRRP in the market, the PRRP relative risks are estimated as a percentage of the excess risk of cigarette smoking.

$$PRRP\ User\ Relative\ Risk = 1 + (Current\ Smoker\ Relative\ Risk - 1) \times PRRP\ Risk\ Ratio$$

Where both products have both been used, either currently or previously, we use the maximum excess risk method. For example, the dual user relative risk (RR)

*Dual User RR = 1 + Max(Current Smoker RR -1, (Current Smoker RR-1) x PRRP Risk Ratio)*

*PRRP User Former Smoker = 1 + Max(Former Smoker RR -1, (Current Smoker RR-1) x PRRP Risk Ratio)*

An independent evidence review published by Public Health England (PHE) (McNeill et al, 2018) concluded that e-cigarettes are 95% less harmful than cigarette smoking, we used this to set the e-cigarette risk ratio to 5%. More recently introduced tobacco heated products which produce nicotine from tobacco but without the combustion—have been estimated to be around 90% less harmful than conventional cigarettes (UK Government Science Review Committee: Reducing Harm – Heat-not-burn tobacco products (para 28), (2018)).

There is lack of agreement about the potential effects of dual use in consumers, which are likely to be diverse depending on many factors. The report by Public Health England (2018) concluded that Dual use is unlikely to be associated with substantial reductions in harm, particularly when there is no substantial reduction in the number of cigarettes smoked, but that comparison between dual users and smokers also indicated that dual use is not associated with an increase in harm.

Mortality rates are calculated from:

$$\text{Never Mortality Probability (Gender, Age)} = \frac{Expected\ Deaths(Gender, Age)}{\sum_i\ Smoking\ Status\ Prevalence(Gender, Age)\ x\ Relative\ Risk(Gender, Age)}$$

where (i) is smoking status (Never, Current, Former), and:

$$\text{Mortality Probability}_{\text{Smoking Status}}\text{ (Gender, Age)} =$$

$$\text{Never Smoker Mortality Probability (Gender, Age)} * \text{Relative Risk (Gender, Age)}_{\text{Smoking Status}}$$

The estimate of the population attributable fraction (PAF) to smoking is calculated, for each gender x age using Levin's formula [Hanley JA, 2001]:

$$PAF = \frac{\sum_i(Prevalence(i) x\ (Relative\ Risk\ (i)-1)}{\sum_i(Prevalence(i) x\ (Relative\ Risk\ (i)-1)+1}$$

Where (*i*) is smoking status : (Never, Current, Former)

and annual smoking attributable deaths are calculated:

$$\text{Smoking Attributable Deaths(Gender, Age)} = \text{PAF(Gender, Age)}\ x\ \text{Total Mortality Rate(Gender, Age)}$$

To estimate Years of Life Lost (YLL) due to smoking, we multiple the smoking attributable mortality rate by the expected period life expectancy remaining by age and sex from ISTAT(c) Life Tables. The YLL is then cumulated over the period of the projection to provide a single statistic.

$$\text{YLL} = \text{Smoking Attributable Deaths(Gender, Age)} * \text{Expected Life Years Remaining(Gender, Age)}$$

Population aging occurs at each time step. As the model time step and age cohort width are both one year, all of the people that remain in a smoking status stock (less those that die) are automatically shifted into the next single year age cohort at each calculation sequence. For those that transition, the stock outflow is at current *age* but then is transformed to *age+1* for the destination inflow.

**E-cigarette Transition Probabilities**

At the date of this paper, a literature search identified no comprehensive e-cigarette transition probabilities studies published for Italy. Rather than make subjective assumptions on the probabilities we applied previously estimated transitions (Brouwer et al. 2020) estimated across 2013–2017. The estimates are for the United States and are based on "The Population Assessment of Tobacco and Health (PATH) Study". The PATH Study is a national longitudinal study of tobacco use and how it affects the health of 49,000 people in the United States.

Brouwer et al 2020 reported initiation probability estimates from Never smokers to:

- Exclusive Cigarette User
- Exclusive electronic nicotine devices (ENDS)
- Dual Use
- Non-Current Use

Two other sources of e-cigarette transition patterns based on data from the PATH Study were identified. (Coleman et al, 2019) and (Wei et al, 2020). We used the estimates from Brouwer et al as they were more detailed, but a comparison of e-cigarette transition estimates from all three sources is provided in the Model Verification supplement to this paper.

<u>E-cigarette Initiation</u>

*Table 5 E-cigarette Initiation Probability*

| Annual Initiation Probability (95% Confidence Intervals) | E-Cigarettes Only | Dual User |
|---|---|---|
| From Never User | 0.3 (0.2 – 0.3) | 0.1 (0.1 – 0.1) |

The estimated e-cigarette initiation probabilities (Table ) are for the entire Never Smoker population across all population ages and gender. The Brouwer paper reported hazard ratios for e-cigarette initiation are:

*Table 61 E-cigarette Initiation Age Hazard Ratios*

| Age | 18-24 | 25-34 | 35-54 | 55+ |
|---|---|---|---|---|
| Hazard Ratio | 27.6 (8.12, 93.9) | 6.8 (1.80, 25.7) | 2.0 (0.52, 7.6) | 1.0 (ref) |

The hazard ratios (Table 61) indicate that e-cigarette initiation occurs at young ages, similar to that observed for smoking initiation. In our model, we calculate the total number of e-cigarette initiators for each year using the total population of Never Smokers, and then distribute those initiators to ages using the same distribution used for cigarette smoking initiation. The same process is applied for Dual User initiation.

The reported initiation probabilities from Never Smoker to Non-Current use were not used. These people were assumed to remain as Never Smokers in the current year, allowing them to potentially initiate at later ages.

E-cigarette Transitions

The e-cigarette transition probabilities used by the model are shown in Table

*Table 7 E-cigarette Transition Probabilities*

| Transition Probability (95% Confidence Intervals) | Cigarettes Only | E-Cigarettes Only | Dual User | Non-Current User |
|---|---|---|---|---|
| Exclusive Cigarette Users | | 1.1 (0.9 – 1.2) | 4.3 (4.0 – 4.6) | *From calibration* |
| Exclusive E-cigarette Users | 7.1 (6.1 – 8.1) | | 14.0 (12.2 – 15.9) | 20.8 (18.2 – 23.3) |
| Dual Users | 45.2 (42.4 – 47.9) | 9.6 (8.0 – 11.1) | | 4.3 (4.0 -4.7) |

As we are comparing PRRP scenarios to a smoking only scenario to reflect the population health impact PRRPs may have against if they did not exist, the quit probability of cigarette smokers remains the same as the probabilities estimated by model calibration.

Former E-cigarette User Relapse

Brouwer et al. (2020) used a combined grouping of Non-Current Users. This could include people that have never smoked or never used an e-cigarette, nor could we distinguish to composition of the group by Former Users of cigarettes, e-cigarettes or both products.

With this limitation, we instead used the same relapse probabilities, based on duration of abstinence, we used for Former Smoker relapse rates. Additionally, we conducted our own analysis of the Path Study data to determine the destination product on relapse (Table )

*Table 8 Relapse Product Destination*

| Relapse Destination | Same Product | Alternative Product | Dual Use |
|---|---|---|---|
| Male | 66.6% | 6.7% | 26.7% |
| Female | 74.2% | 16.1% | 9.7% |

**THP Transition Probabilities**

THP transition probabilities are not available yet for Italy. A published report of the results of a cross-sectional epidemiological pilot survey of THP use in Japan (Adamson et al. 2020) and (Camacho et al. 2021) and a larger follow-up study currently under review in Tobacco Regulatory Science were used as substitute values.

There is a difference in definition of smoking status in this report. A Current User (smoker or THP) is defined as someone who has used the product 100+ times in their lifetime and last used within 12 months. A Former User is someone that has used the product 100+ times in their lifetime and not used within 12 months.

THP Initiation

The THP initiation probabilities used by the model are shown in Table 9

*Table 9 THP Initiation Probability*

| Annual Initiation Probability (95% Confidence Intervals) | THP Only | Dual User |
|---|---|---|
| From Never User | 0.1% (0.0 – 0.4) | 0.0% (0.0 – 0.1) |

Again, as with e-cigarette initiation probabilities, these probabilities are based on the entire Never Smoker population. In our model, we calculate the total number THP initiators for each year using the total population of Never Smokers, and then distribute those initiators to ages using the same distribution used for cigarette smoking initiation. The same process is applied for Dual User initiation.

THP Transitions

The THP transition probabilities used by the model are shown in Table

*Table 10 THP Transitions*

| Transition Probability (95% Confidence Intervals) | Cigarettes Only | THP Only | Dual User | Non-Current User |
|---|---|---|---|---|
| Exclusive Cigarettes Users | | 0.6% (0.2 – 1.3) | 9.5% (7.6 – 11.7) | *From calibration* |
| Exclusive THP Users | 0.0 | | 3.0% (0 – 20.5) | 5.1% (0.8 – 15.9) |
| Dual Users | 4.6% (1.8 – 9.2) | 32.8% (25.7 – 40.4) | | 2.1% (0.5 – 5.9) |

As we are comparing PRRP scenarios to a smoking only scenario to reflect the population health impact PRRPs may have against if they did not exist, the quit probability of cigarette smokers remains the same as the probabilities estimated by model calibration.

**Demographic Inputs**

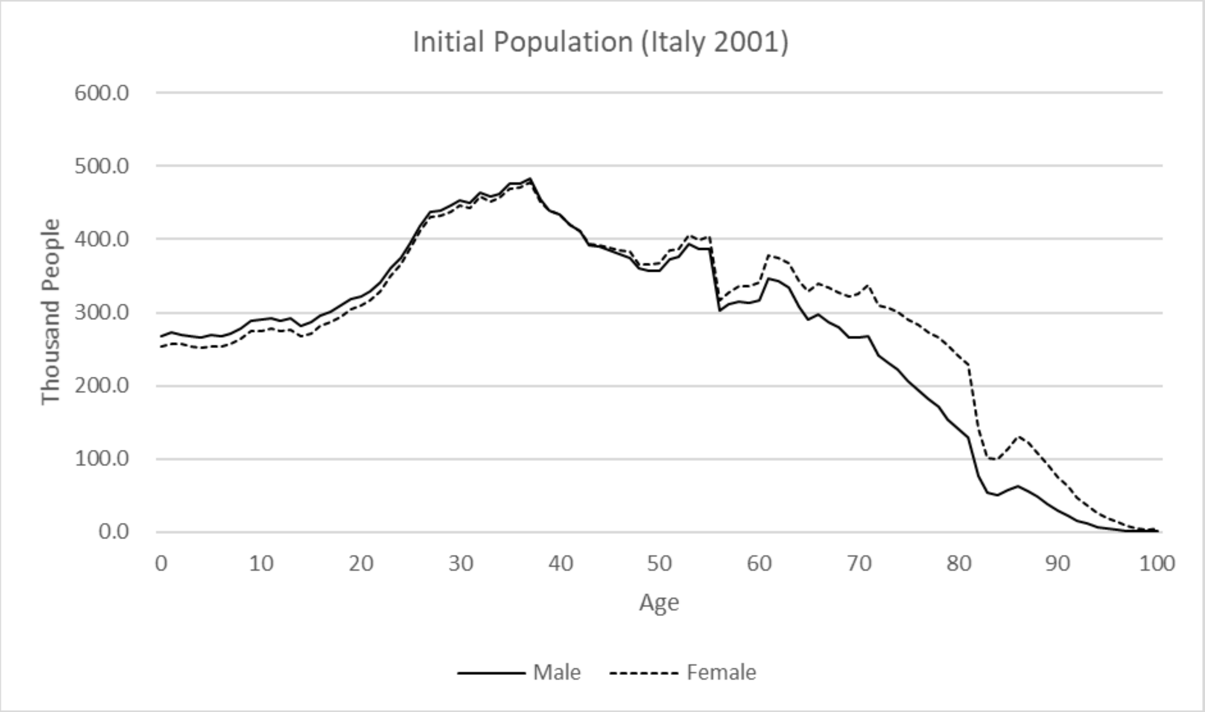


*Figure 3 Italian Population by Gender and Age (2001)*

Data Source: ISTAT - Resident Population on 1st January

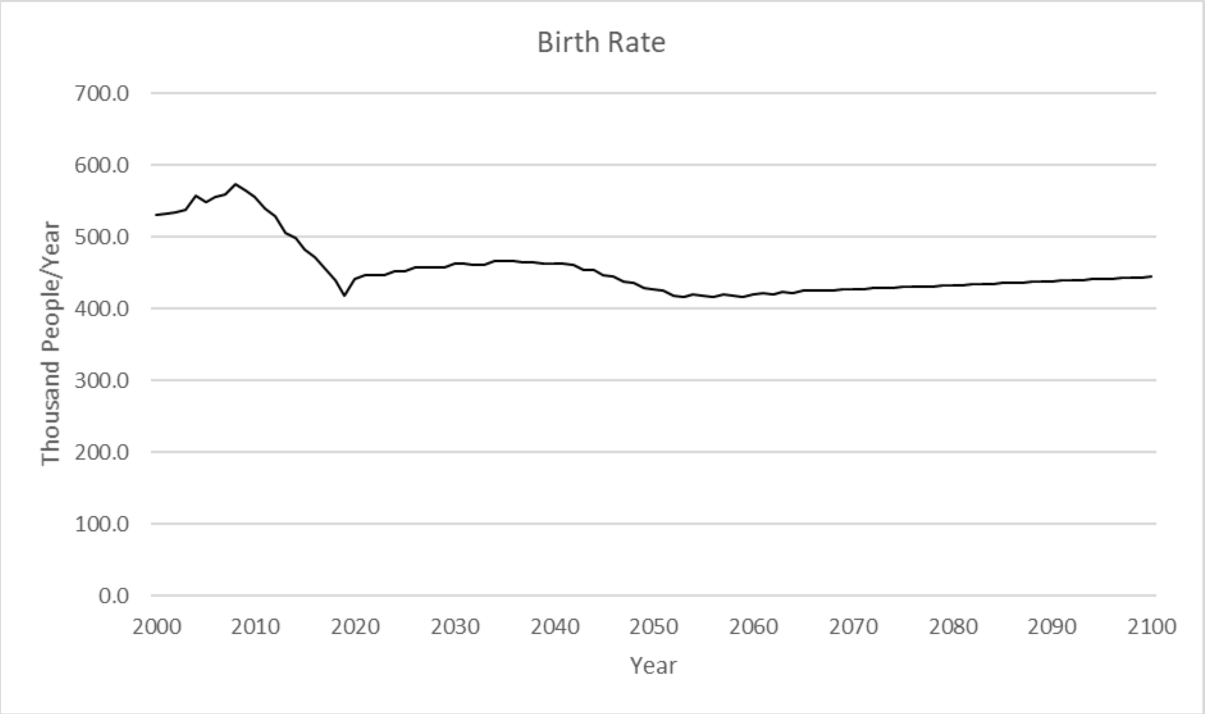


*Figure 4 Projected Italian Birth Rate*

Data Source: ISTAT(b) – Population Projection (2018-2065)

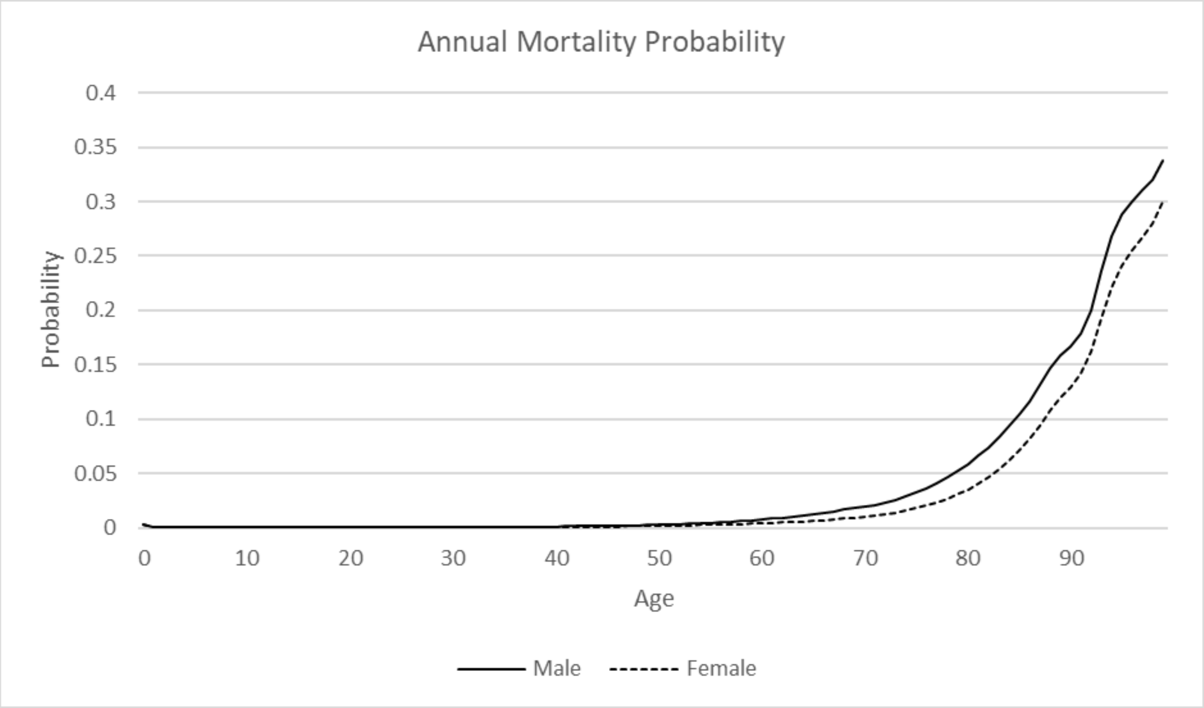


*Figure 5 Mortality Rate (qx) by Gender and Age*

Data Source: ISTAT(c) Life Tables (2012)

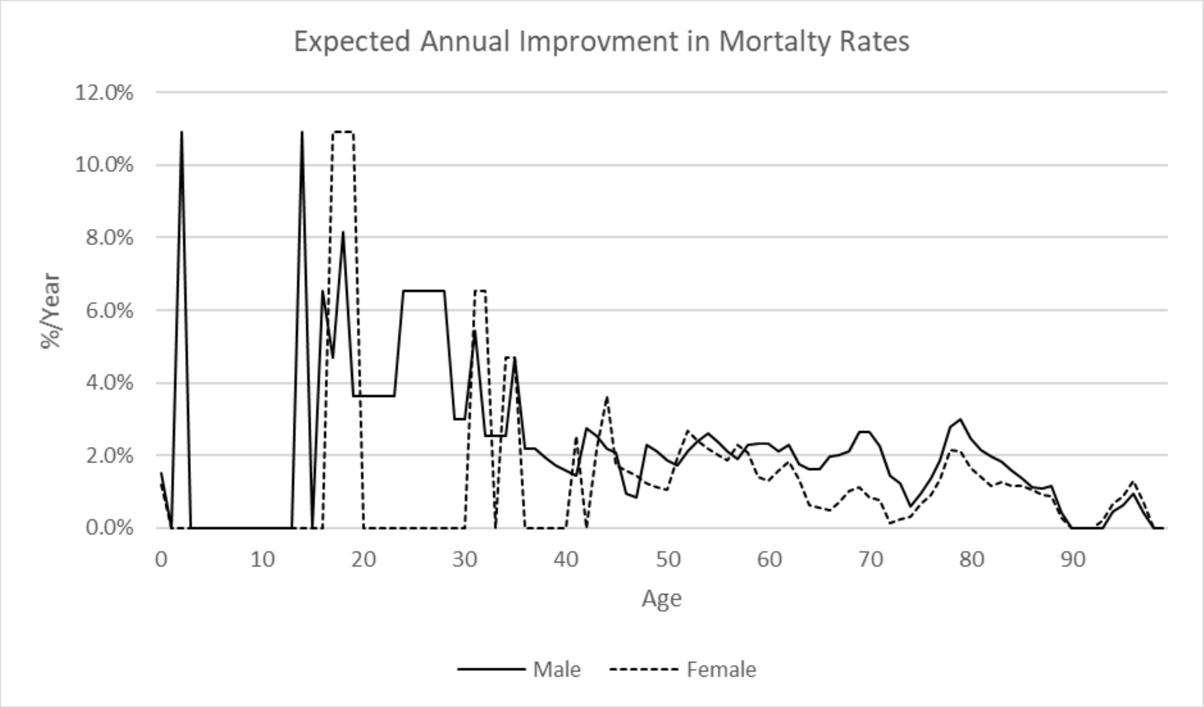


*Figure 6 Expected Annual Improvement in Mortality Rates for Gender and Age*

Data Source: ISTAT(c) Life Tables (2012)

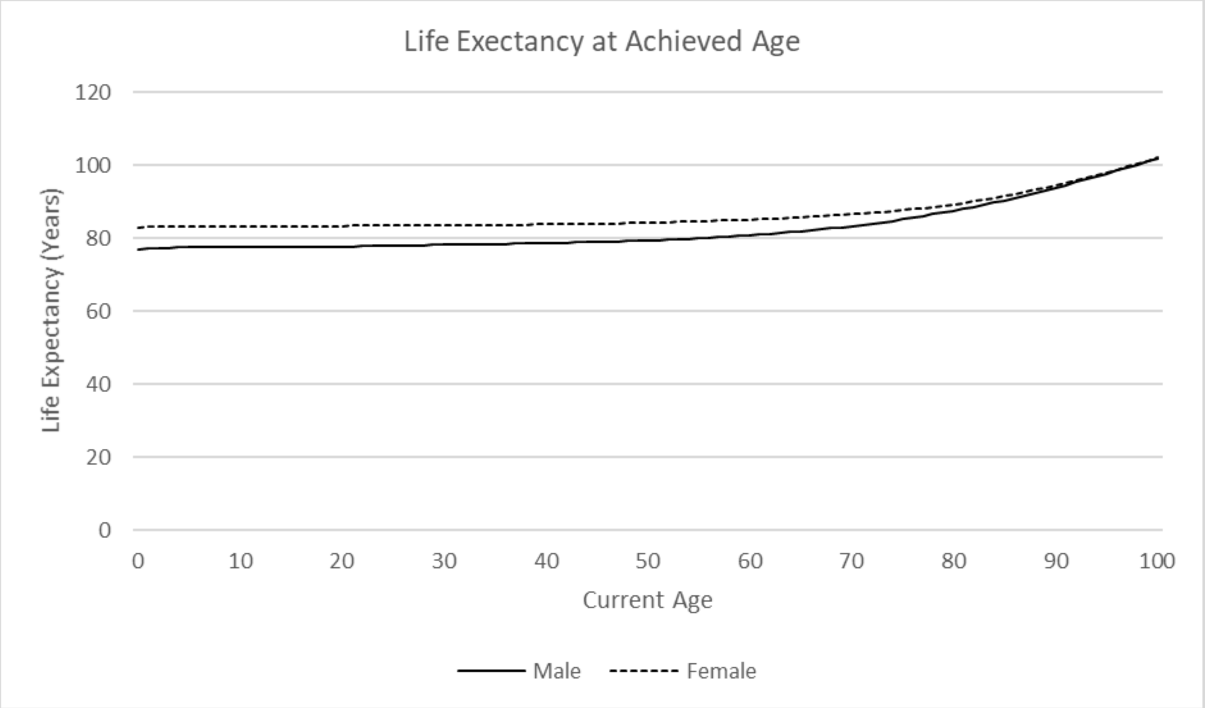


*Figure 7 Life Expectancy by Gender and Current Age*

Data Source: ISTAT(c) Life Tables (2012)

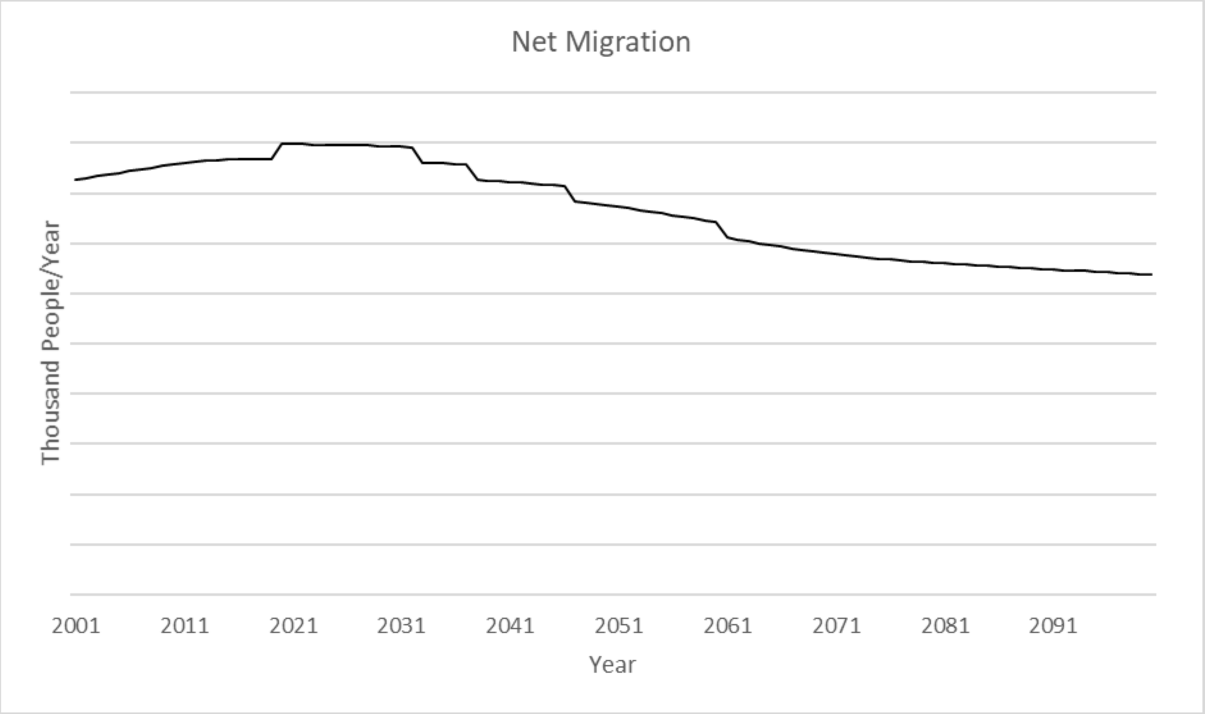


*Figure 82 Projected Net migration*

Data Source: ISTAT(b) – Population Projection (2018-2065)

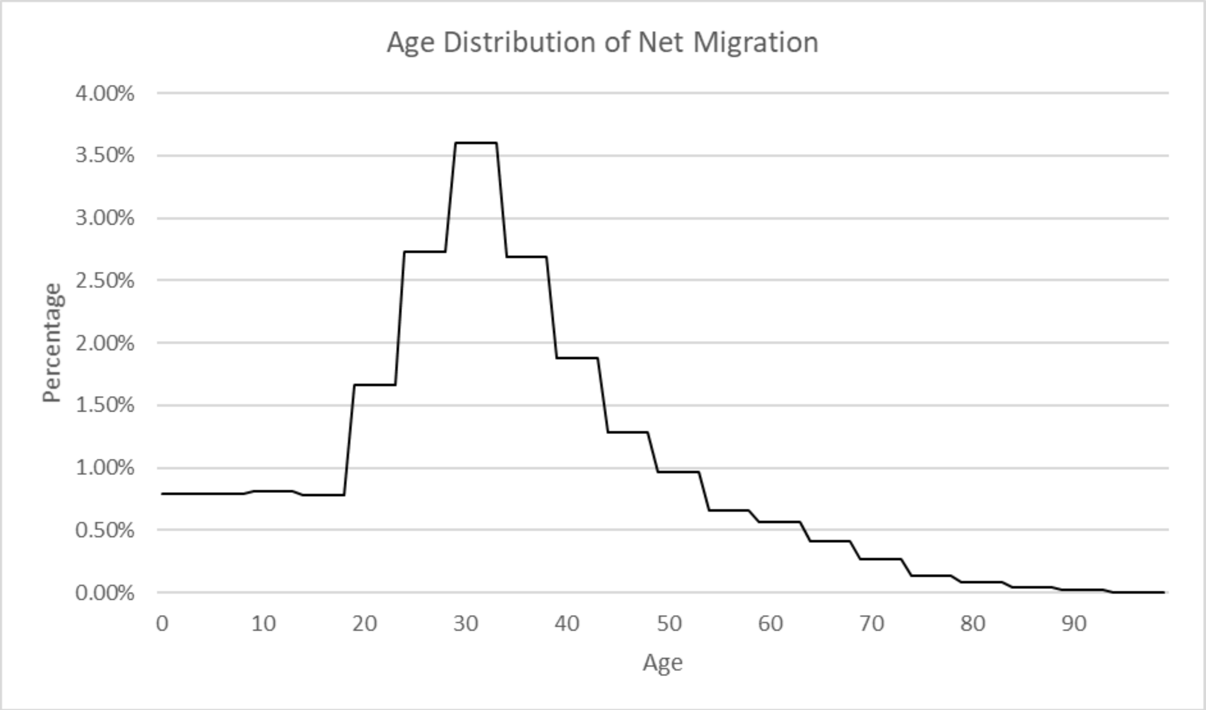


*Figure 9 Net Migration Age Demographics*

Data Source: EUROSTAT (b) – (Net Migration(2020)/Population(2020) by gender and age)

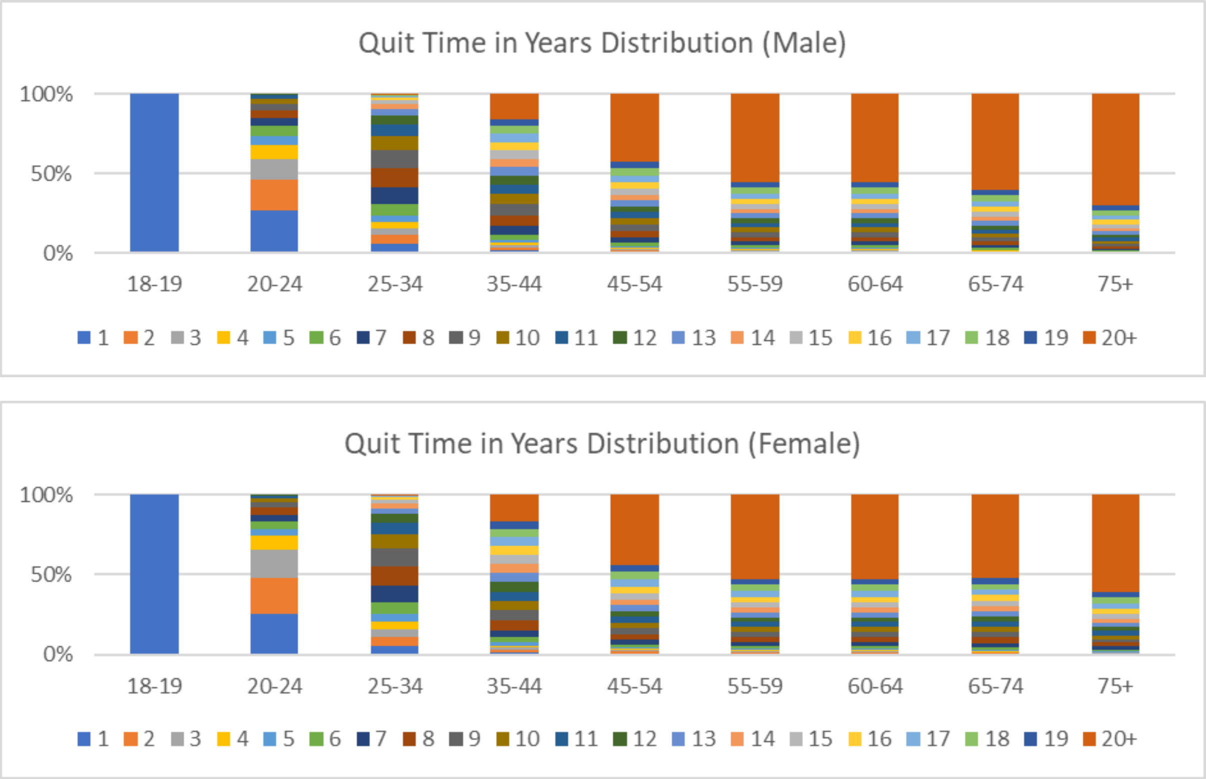


*Figure 10 Former Smoker Quit Time Distribution*

Data Source: Population Assessment of Tobacco and Health (PATH) Study (US DHHS 2020)

### Model Assumptions

*Smoking Initiation*

No smoking initiation under the age 14 is assumed.

*PRRP Transition Probabilities*

E-cigarette transition probability estimates from a sample of the United States population (Brouwer et al, 2020) and THP transitions from Japan (Adamson et al,2018) and (Camacho et al. 2021) are used as substitutes for the Italian population. There will be differences in smoking behaviour across countries influenced by factors such as varying legal smoking ages, taxation levels and smoking restrictions. However, without detailed Italian studies available, assuming alternative country data was concluded as a better approach than basing PRRP transition rates purely via suppositions.

*Relapse*

As e-cigarettes and THPs have only been in the market for a decade, long term relapse data and studies are not yet available. In addition, the e-cigarette transitions from Brouwer et al (2020) grouped all Former Product users together as "Non Current Users" of tobacco products. This precluded us from distinguishing whether transitions from "Non Current Users" were relapses back to current use of the same product or to an alternative products. In the scenarios projected we assume the same relapse probability curve as used for former smokers for both former e-cigarette and THP users.

*Proportion of PRRP initiation that would have smoked.*

Cigarette initiation probabilities are estimated prior to significant PRRP prevalence. It has been argued that it is the risk-taking nature of a person that drives the causal effect of whether they initiate a tobacco product (Kim & Selya. 2020). The people that take up e-cigarettes or THPs from never tobacco status are the same kind of people that would have taken up smoking had alternative products not been available. We use an assumption that 50% of people initiating a PRRP would have initiated cigarettes had the PRRP not been available and adjust the smoking initiation flow accordingly.

*Net Migration Smoking Status*

Net migration flows are assumed to have the same tobacco use status distribution as the Italian population at the time. Distributions are applied at the gender and single year age cohorts.

**Model Outputs**

All model variables are available as outputs. A selection of key output indicators is shown below.

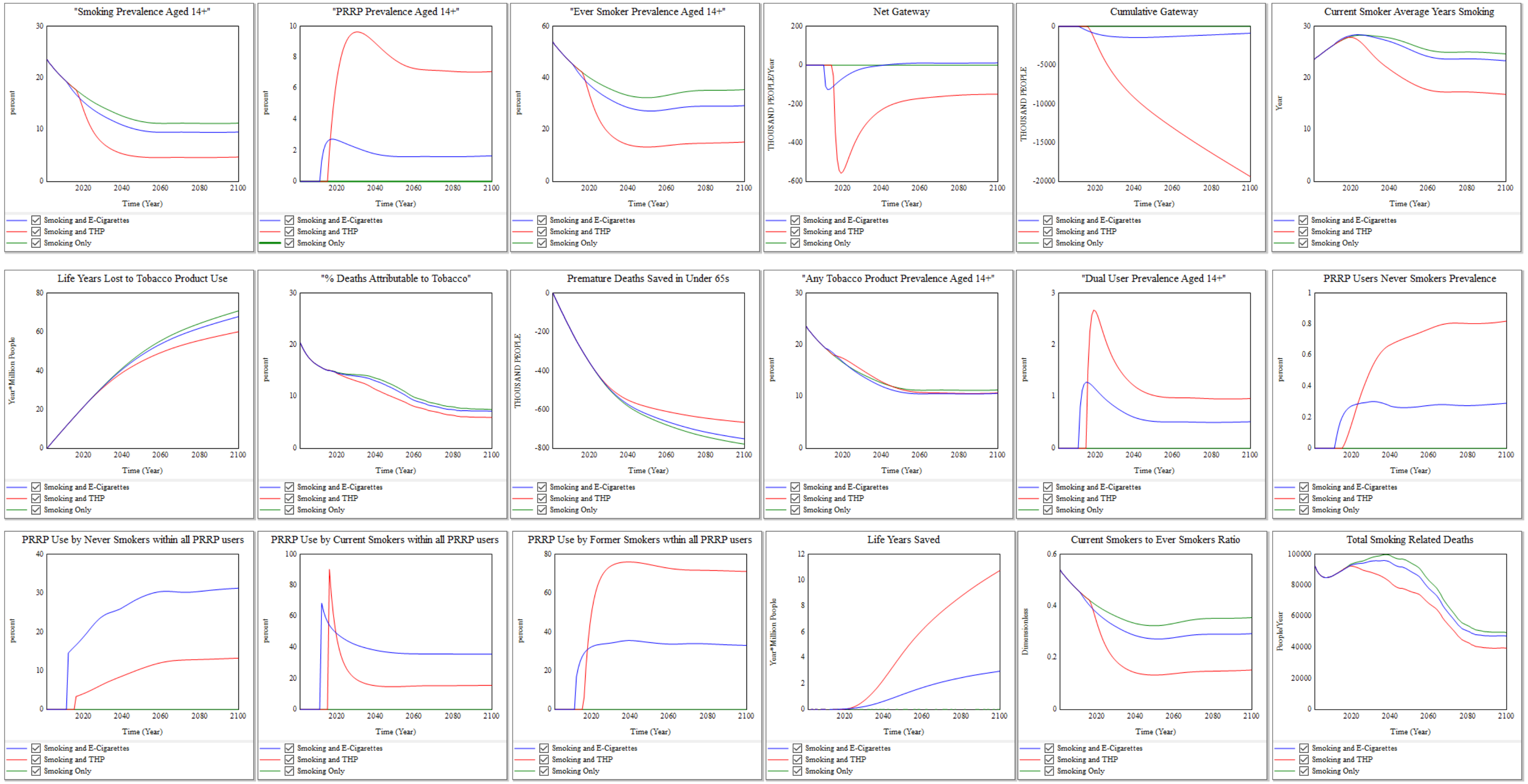

"Smoking Prevalence Aged 14+"
"PRRP Prevalence Aged 14+"
"Ever Smoker Prevalence Aged 14+"
Net Gateway
Cumulative Gateway
Current Smoker Average Years Smoking
Life Years Lost to Tobacco Product Use
"% Deaths Attributable to Tobacco"
Premature Deaths Saved in Under 65s
"Any Tobacco Product Prevalence Aged 14+"
"Dual User Prevalence Aged 14+"
PRRP Users Never Smokers Prevalence
PRRP Use by Never Smokers within all PRRP users
PRRP Use by Current Smokers within all PRRP users
PRRP Use by Former Smokers within all PRRP users
Life Years Saved
Current Smokers to Ever Smokers Ratio
Total Smoking Related Deaths
Time (Year)
Smoking and E-Cigarettes
Smoking and THP
Smoking Only
