## Supplementary material for "Investigating the health effects of 3 coexisting tobacco and nicotine products using system dynamics population modeling: An Italian population case study": 2-product Model Verification Final.pdf

***Supplementary File 3 -Two-Product Model Verification***

*Dimensional Consistency*

All model variables were assigned unit of measurement. All model equations passed the Vensim software dimensional consistency test.

*Demographic Projections*

The model demographic projection of the total population of Italy and the average age of inhabitant's closely matches the National Population Projection 2018-2065 (ISTAT (a) – Demographic Projection).

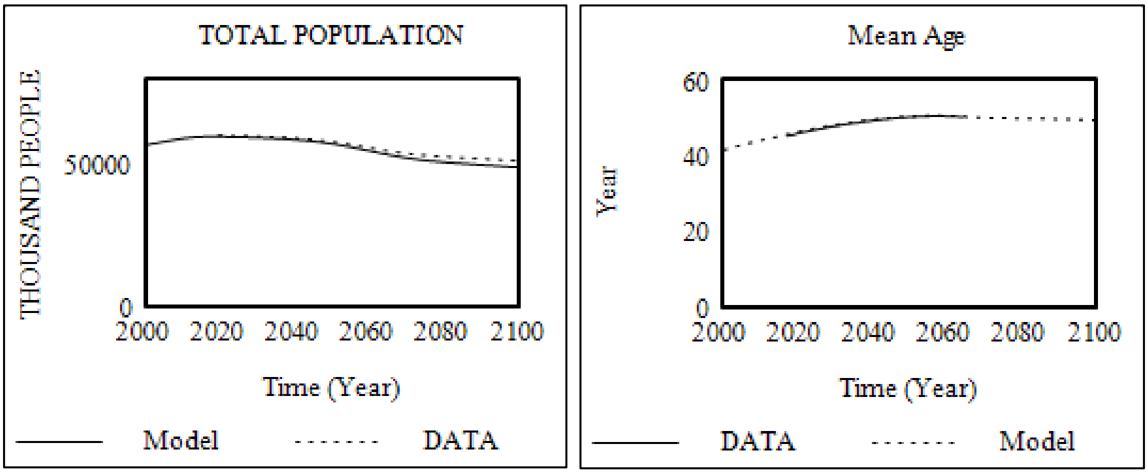


*Figure 1 Population Projection Comparisons*

*Calibrated Smoking Prevalence*

The aggregated prevalence of Never, Current and Former smoking prevalence for those aged 14+ closely aligned to the estimated reported by ISTAT over the same period.

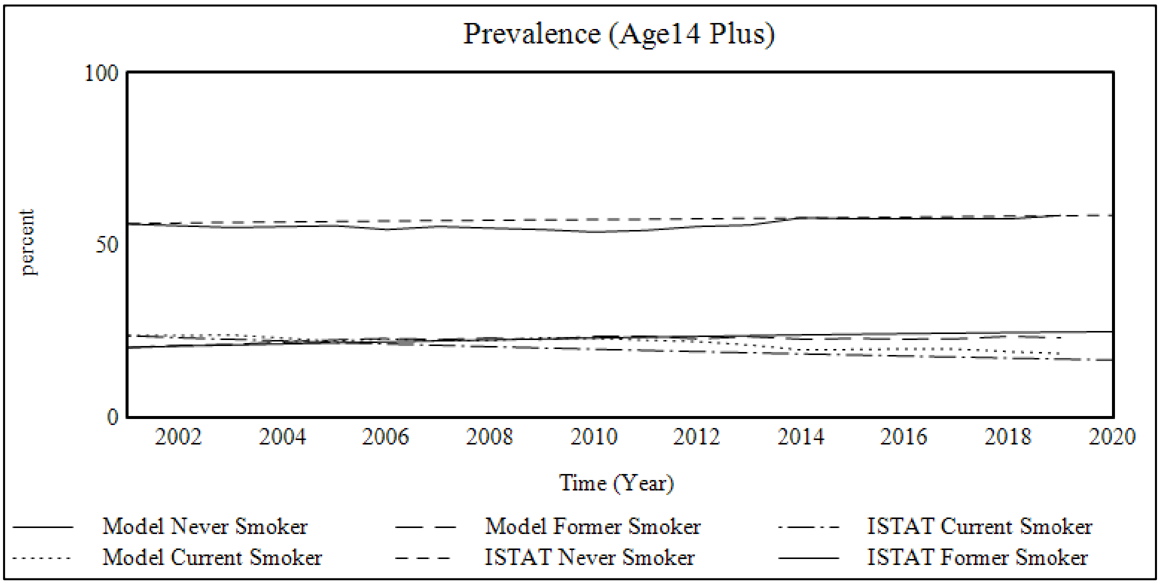

Prevalence (Age14 Plus)
100
50
0
percent
2002
2004
2006
2008
2010
2012
2014
2016
2018
2020
Time (Year)
Model Never Smoker
Model Former Smoker
ISTAT Current Smoker
Model Current Smoker
ISTAT Never Smoker
ISTAT Former Smoker

*Male Smoking Calibrated Prevalence*

*Age 14-17*

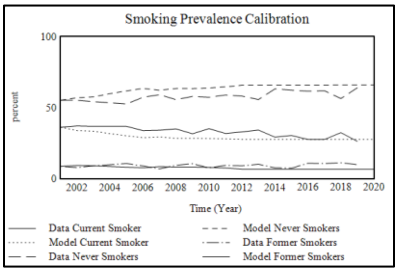


*Age 18-19*

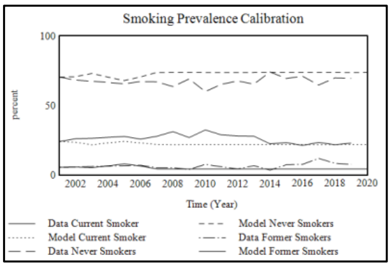


*Age 20-24*

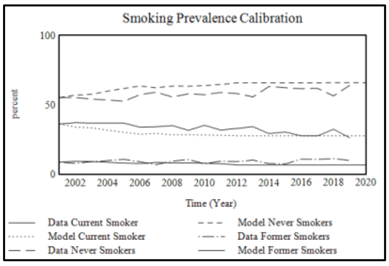


*Age 25-34*

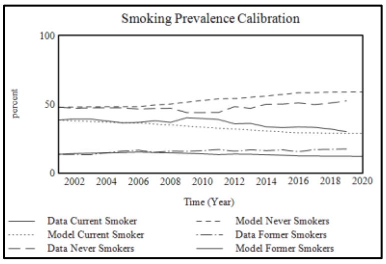


*Age 35-44*

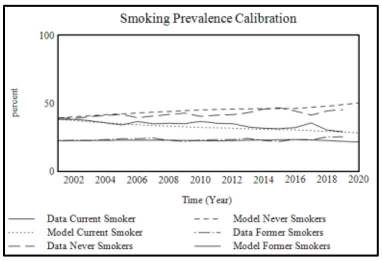


*Age 45-54*

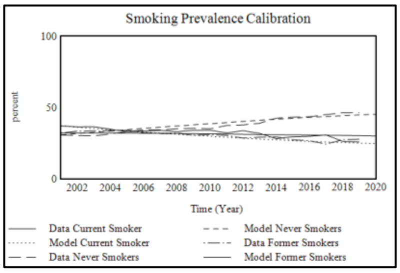


*Age 55-59*

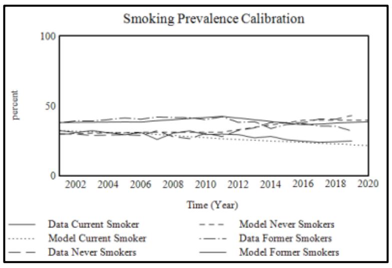


*Age 60-64*

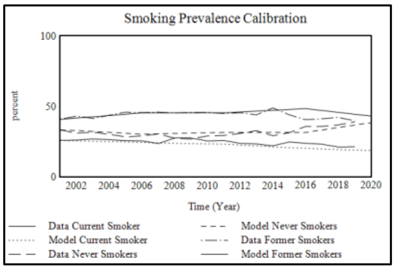


*Age 65-74*

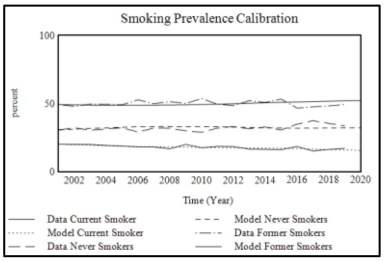


*Age 75+*

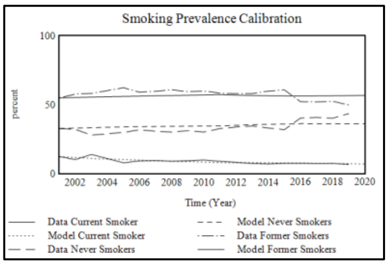

*Female Smoking Calibrated Prevalence*

*Age 14-17*

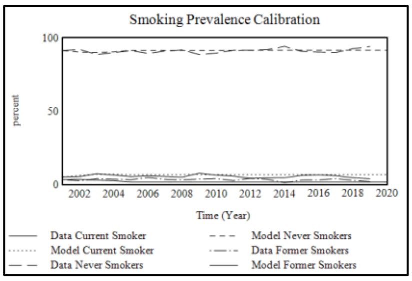


*Age 18-19*

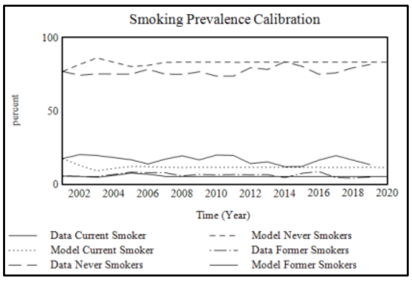


*Age 20-24*

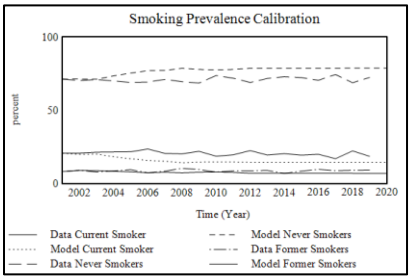


*Age 25-34*

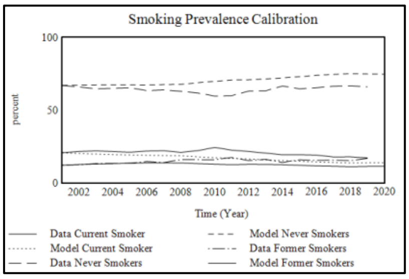


*Age 35-44*

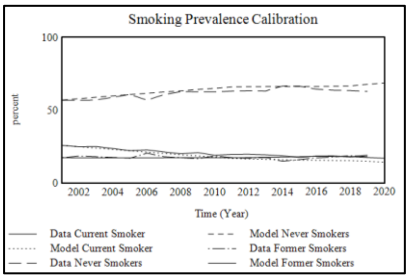


*Age 45-54*

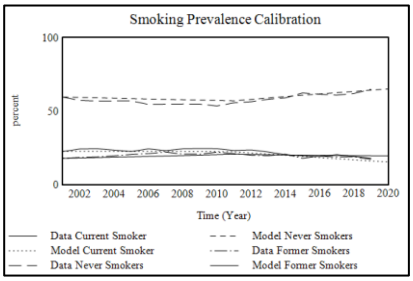


*Age 55-59*

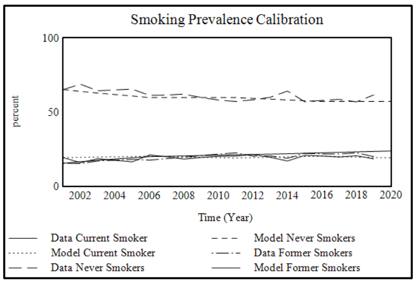


*Age 60-64*

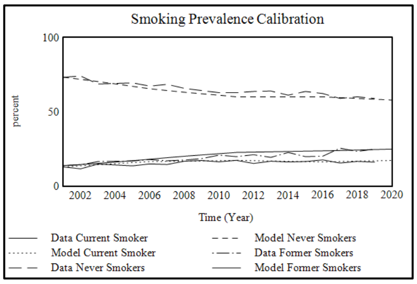


*Age 65-74*

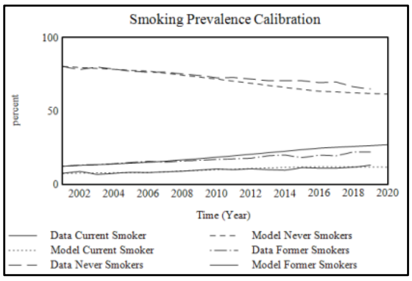


*Age 75+*

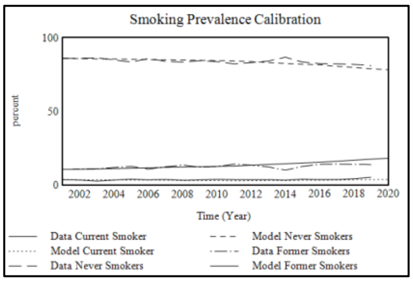

*Smoking Attributable Mortality*

The model estimation of the percentage of deaths attributable to smoking closely aligned to the estimated share of premature deaths attributed to tobacco smoking reported in the annual Global Burden of Disease Report (GBDR, 2019).

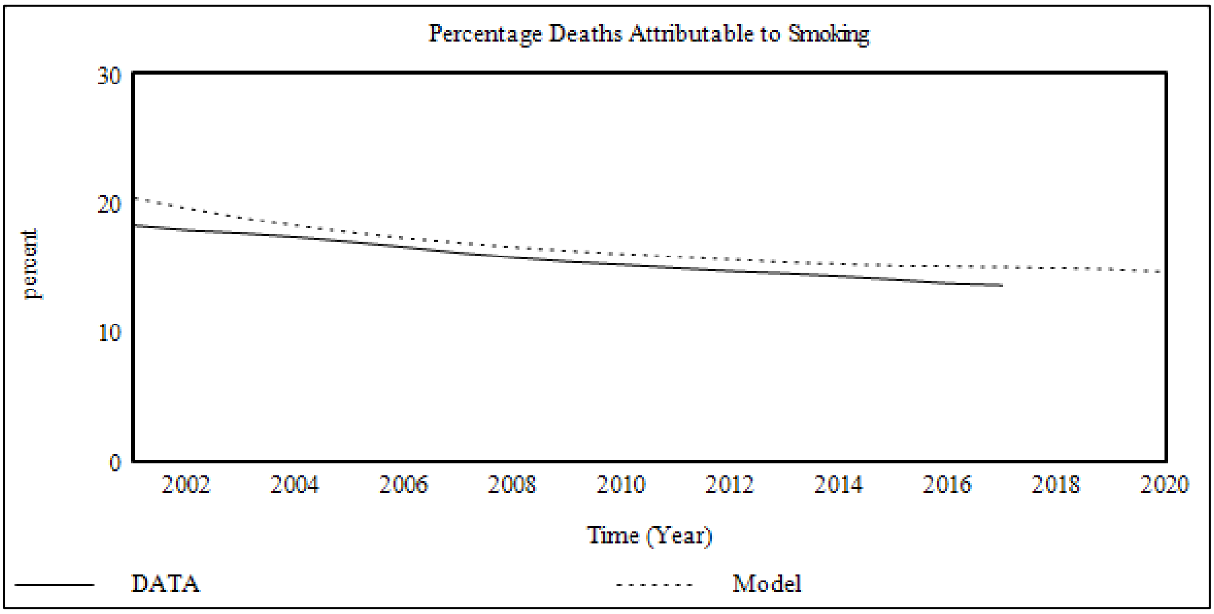


*Mass Balance Check (MBC)*

As the model is formed as a closed system a Mass balance equation was added to check for any leakage to or from the closed system. The MBC error did not exceed 10-6 thousand people at any time point.

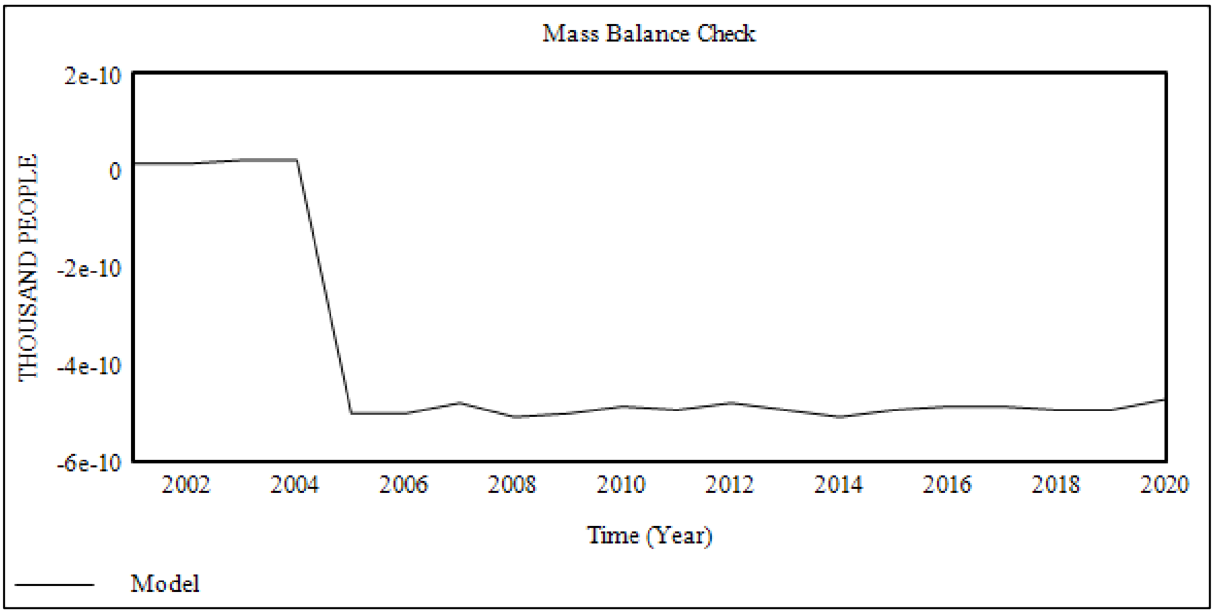


*Extreme Values Robustness*

Using a multivariate sensitivity testing all transition probabilities were set to be randomly selected between 0% to 100% with an interval of 100%. Model calculated prevalence of all products were checked to be within the percentage range 0-100 and that the Mass Balance Check equation for a closed system was still true.

*E-Cigarette Transition Probabilities*

The transition probabilities used in the model from the University of Michigan are published in the BMJ Journal – Tobacco Control (Brouwer et al. 2020). These were compared to an alternative source estimates (Wei et al. 2020) to check for consistency in probability estimates. Both studies used raw data from the PATH Study but using different number of survey waves. Similar patterns of transition behaviour were found in both studies.

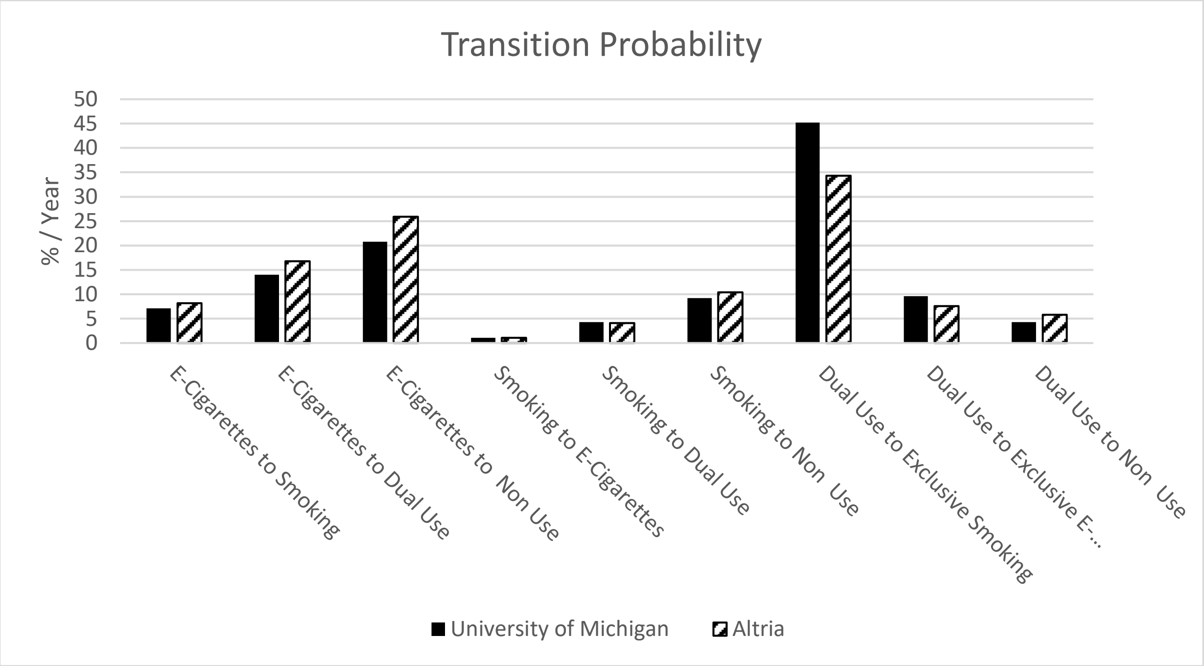


Although studies are available on THP prevalence (Tabuchi et al. 2018), no other study was identified that could provide a comparison to the model THP transition probabilities.

*<u>Additional sensitivity outputs</u>*

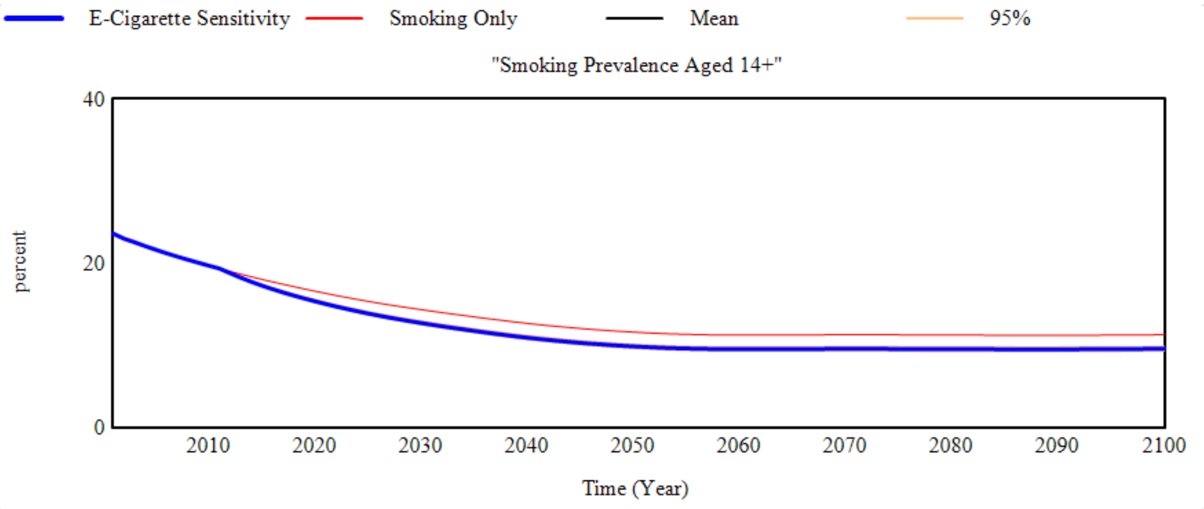


*Figure 2. Smoking prevalence sensitivity to e-cigarette transition probabilities in the E-Cigarette scenario.*

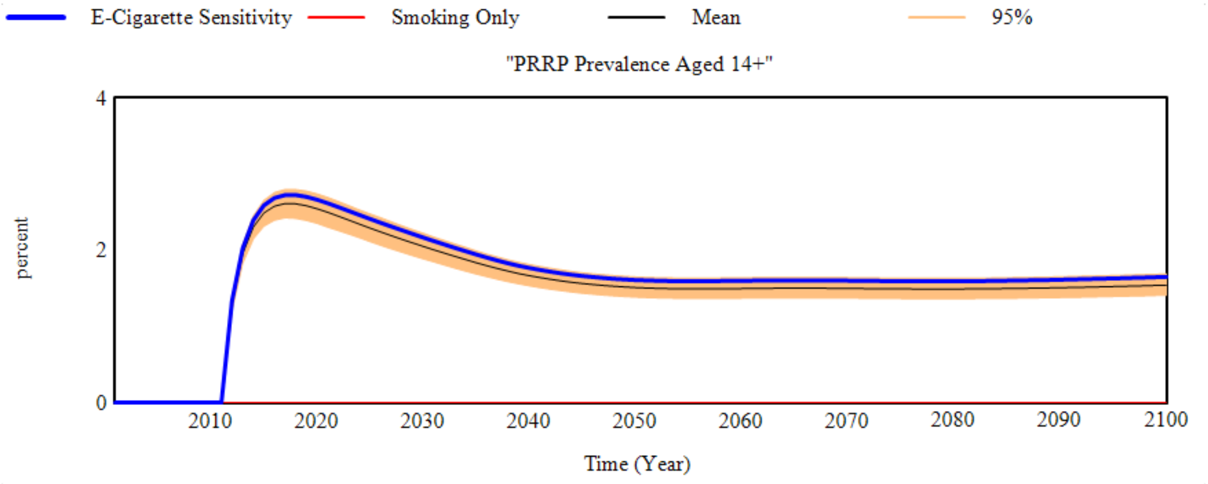


*Figure 3. E-Cigarette prevalence sensitivity to e-cigarette transition probabilities in the E-Cigarette scenario.*

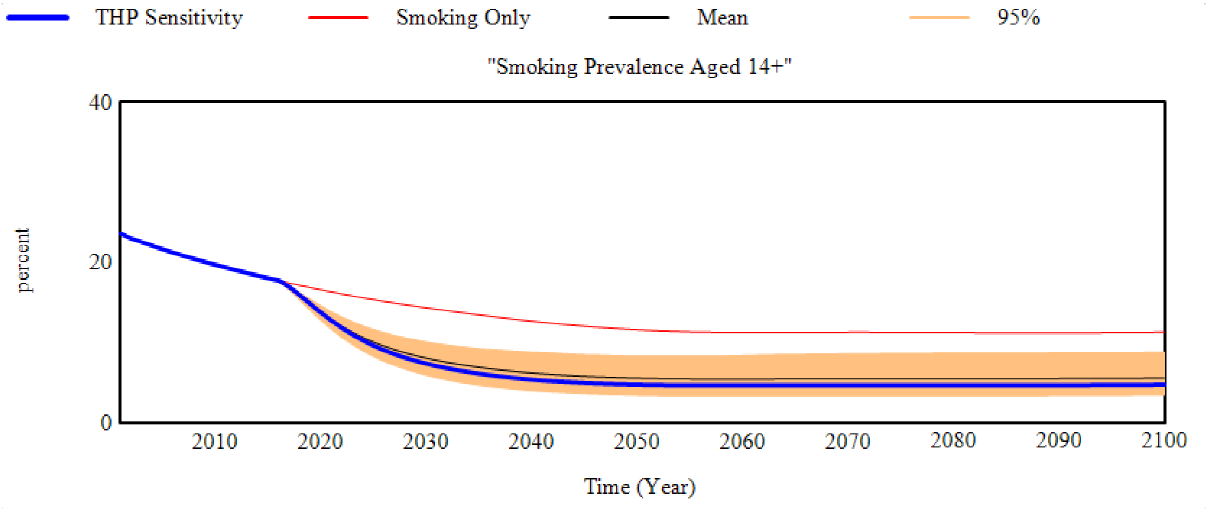


*Figure 4. Smoking prevalence sensitivity to THP transition probabilities in the THP scenario.*

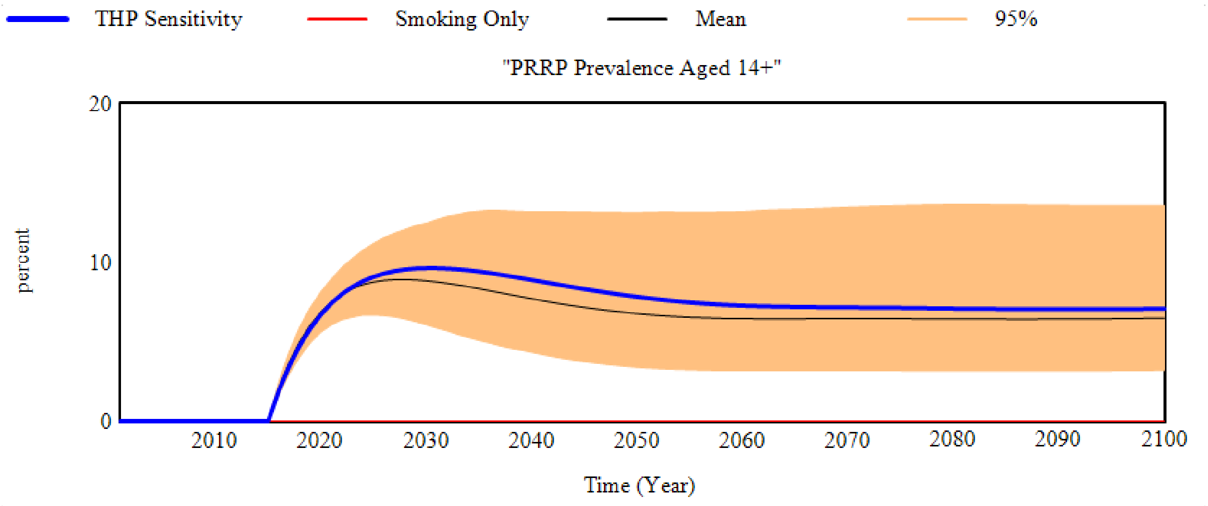


*Figure 5. THP prevalence sensitivity to THP transition probabilities in the THP scenario.*
